# Impact of the training, support and access model (TSAM) on patient health outcomes in Rwanda: controlled longitudinal study

**DOI:** 10.1101/2024.10.24.24316071

**Authors:** Celestin Hategeka, Larry D Lynd, Cynthia Kenyon, Anaclet Ngabonzima, Isaac Luginaah, David Cechetto, Michael R Law

## Abstract

Achieving maternal and newborn health (MNH) related Sustainable Development Goal targets will require high-quality health systems in low– and middle-income countries. While over 90% of deliveries in Rwanda take place in health facilities, maternal and neonatal mortality rates remain high. In an effort to bolster quality of care provided to women and newborns to ultimately reduce morbidity and mortality, the Training, Support and Access Model (TSAM) clinical mentorship was established in 10 district hospitals in Rwanda in 2017. We evaluated the impact of the TSAM clinical mentorship intervention on maternal and newborn health outcomes. We used monthly time series data from the DHIS2-enabled Rwanda health management information system from February 2014 to February 2020 to assess the impact of the TSAM intervention on outcomes of care for MNH in intervention hospitals relative to concurrent control hospitals. Using a controlled quasi-experimental interrupted time series analysis, we estimated changes in rates of inpatient mortality and morbidity for MNH associated with the implementation of the TSAM clinical mentorship. The study cohort included 25 hospitals (10 TSAM hospitals and 15 control hospitals) that collectively reported 339,850 hospital deliveries and 94,584 neonatal hospital admissions. We found that the implementation of the TSAM clinical mentorship intervention was associated with a two-years reduction of 84% in the obstetrical complication case fatality rate, 32% in hospital neonatal mortality rate, 30% in postpartum hemorrhage incidence rate, and 48% in neonatal asphyxia incidence rate in TSAM hospitals relative to control hospitals. However, the stillbirth rate did not decline following the TSAM intervention. We found that a quality improvement strategy that employed continuous quality improvement approaches using onsite clinical mentorship of health providers along with involvement of health facility leadership to facilitate the improvement was associated with improvements in MNH in Rwanda. Our findings provide evidence that can justify the scale up of TSAM across the country and potentially in other similar settings.

**Summary box:** *What is already known?:*

- Poor quality of healthcare is currently a bigger driver of excess maternal and neonatal mortality than under-utilization of health facilities in many low– and middle-income countries (LMICs).
- Achieving maternal and newborn health related Sustainable Development Goal targets will require high-quality health systems in LMICs.

*What does this study add?:*

- The Training, Support and Access Model (TSAM) clinical mentorship implemented in 10 Rwandan district hospitals to bolster quality of care provided to women and newborns was associated with a reduction in in-hospital maternal and newborn deaths. However, the (intrapartum) stillbirth rate did not decline following the TSAM intervention.
- The TSAM intervention was associated with a significant decline in in-hospital maternal and neonatal morbidity (e.g., incidence of postpartum hemorrhage and neonatal asphyxia).

*What do the new findings imply?:*

- Employing continuous quality improvement approaches using onsite clinical mentorship of health providers along with involvement of health facility leadership to facilitate the improvement can be an effective strategy to improve maternal and newborn health outcomes.
- Quasi-experimental methods leveraging routine health information systems data can be useful to study impact of health system improvement interventions in low-resource settings.

## 1. Introduction

Improving maternal and newborn health (MNH) outcomes remains a significant challenge in low and middle-income countries (LMICs), which account for over 90% of the global burden of maternal and neonatal mortality.^1, 2^ Bolstering both utilization and quality of currently recommended evidence-based MNH services has increasingly been advocated as a vital strategy to optimize MNH outcomes.^3–5^ While utilization of MNH services has increased considerably over the past few decades, quality of MNH services has lagged behind in many LMICs.^3^ Consequently, using health facilities including for childbirth does not necessarily translate to improved MNH outcomes and, in fact, poor quality of healthcare is currently a bigger driver of excess maternal and neonatal mortality than under-utilization of health facilities in these settings.^6–8^ Similarly, they are not likely to achieve maternal, newborn and child health related Sustainable Development Goal (SDG) targets by 2030 if the current rates of reduction in maternal, neonatal and under-five mortality do not increase.^9^

Mounting evidence suggests that high-quality health systems are needed to accelerate rates of decline in maternal and newborn mortality and ultimately help LMICs achieve MNH related SDG and national ambitions. ^3^ Research suggests that health system quality improvement strategies in these settings have largely employed micro-level interventions (e.g., short term in-service training and / or supervision of healthcare providers) in an attempt to improve quality of care for mothers and newborns.^3^ However, such interventions alone have limited impact without system-level changes targeting quality improvement.^3, 10, 11^ Additionally, system-level changes such as reforming medical education and health service delivery reorganization generally take longer to implement and / or take effect. Therefore, there is an urgent need of strategies that can help bolster quality of care for mothers and newborns while system level changes are being contemplated and / or implemented.

In response, the Training, Support, and Access Model (TSAM) project established a clinical mentorship program in district hospitals in Rwanda in 2017 to contribute to building the capacity of health providers and supporting continuous quality improvement to deliver high quality of care to women, newborns and children. While several other district hospitals (first-line referral hospitals in each of Rwanda’s 30 districts) have either implemented or proposed similar mentorship interventions, there have been conspicuously few rigorous evaluations of the impact of these interventions. Studies that have evaluated the effectiveness of mentorship interventions for health providers suggest that they are generally effective in improving health providers’ performance and processes of care quality; however, there is a paucity of evidence on their impact on outcomes of care (e.g., maternal and perinatal outcomes and under-five mortality) in LMICs.^12–16^ Thus, the objective of this study was to evaluate the impact of the TSAM clinical mentorship intervention on outcomes of care for mothers and newborns in Rwanda.

## 2. Methods

### 2.1. Study context

Rwanda is a low-income country with over 12 million population. It is one of the countries where coverage of many MNH interventions has increased considerably; however, quality of MNH services has not improved as quickly.^6, 17–26^ The country has embarked on system-level changes including reforming medical education through the Rwanda Human Resources for Health program to produce healthcare providers (including specialist doctors and nurses/midwives) with advanced skills necessary to provide advanced care including for MNH emergencies occurring in district hospitals.^27^ While considerable efforts have been deployed to increase the number of specialist health providers deployed in Rwandan district hospitals, many of these hospitals are still staffed by non-specialist health providers who often handle MNH emergencies.^27, 28^ Implementing short-term in-service trainings for healthcare providers in Rwandan district hospitals has largely been hampered by multiple factors including lack of mentorship and refresher trainings, and high staff turnover.^29–31^

To address these challenges and ultimately reduce and sustain a significant impact on mortality and morbidity for maternal, newborn, and child health, it was necessary to develop an integrated, harmonized comprehensive emergency care training accompanied by mentoring, coaching and outreach program to support those trained. In 2015, Global Affairs Canada provided funding to expand the Maternal Newborn and Child health in Rwanda project that had focused on providing training programs to the TSAM program for maternal, newborn, and child health in Rwanda and Burundi. However, because of conflicts that were ongoing in Burundi, the project was only implemented in Rwanda.

### 2.2. Study intervention

In collaboration with the Rwanda Ministry of Health, the TSAM project established a clinical mentorship program in 10 district hospitals in Rwanda (five in Northern Province and five in Southern Province) based on the Ministry of Health recommendation. The mentorship program was designed to build the capacity of health care providers in delivering high quality of care to women, newborns, and children to reduce morbidity and mortality. The TSAM mentorship model was built on an inter-professional collaboration approach with a team of five mentors working together in three areas of competency: maternity services including a gynecologist/obstetrician and a midwife; pediatrics services including a pediatrician and a pediatric nurse; and anesthesia including an anesthesiologist physician or a non-physician anesthetist.^32^

The mentors visited their assigned hospitals bimonthly to work with mentees who were also trained in emergency obstetrical, neonatal, and pediatric care. Mentees were trained in continuous quality improvement and were supported in this effort. Each visit lasted for three days. There was a mentee logbook that was used to track performance and progress of mentees until they graduated, typically in one year. The mentorship started in June 2017 in five district hospitals in the Northern Province, and subsequently expanded to five hospitals in the Southern Province in November 2017 with modification to the model to incorporate lessons learnt in North (from June-November 2017) (Figure 1). This TSAM clinical mentorship also included regular monitoring and evaluation meetings where lessons learnt, challenges and experiences were regularly shared with everyone involved in the mentorship including hospital managers (e.g., Director Generals, Clinical Directors and Directors of Nursing) of the intervention hospitals to continuously improve implementation fidelity of the intervention. Overall, the TSAM clinical mentorship activities focused on improving maternal and newborn health outcomes. We have described the TSAM clinical mentorship model development and implementation elsewhere.^32^ All TSAM clinical mentorship activities stopped in March 2020 amidst the country lockdown because of the COVID-19 pandemic.

**Figure 1.**
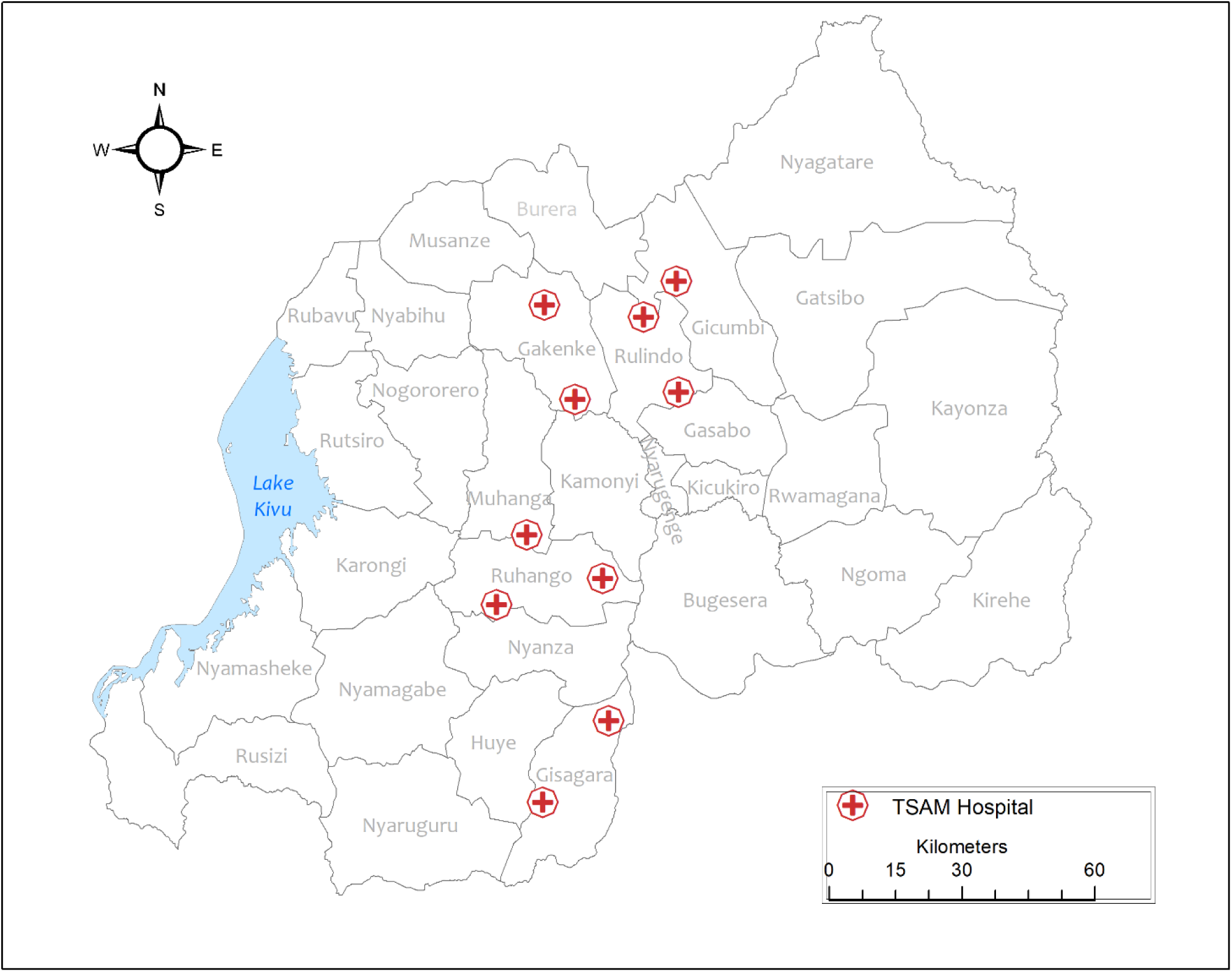
Rwandan hospitals with the TSAM clinical mentorship program.

### 2.3. Data sources

We used time series data from the DHIS2-enabled Rwanda health management information system (Rwanda HMIS) to evaluate the impact of the TSAM clinical mentorship intervention on outcomes of care for MNCH in intervention facilities relative to concurrent controls. Each hospital provided data for 40 months before the intervention and 28 months after the start of the TSAM clinical mentorship. The Rwanda HMIS is a routine health information system which captures health information including on health care utilization, morbidity, and mortality across Rwandan health facilities.^33^ Data are aggregated at facility level and reported each month. Efforts have been made to ensure the quality of the Rwanda HMIS data, including regular data quality checks and audits to ensure data quality.^33^ Previous research assessing the quality of the Rwanda HMIS data found that MNH indicators in the Rwanda HMIS performed well in terms of timeliness, completeness, and consistency.^34, 35^ Similarly, Rwanda HMIS data has been used in prior studies.^10, 34, 36, 37^

### 2.4. Study cohort

We examined the impact of the TSAM intervention on health outcomes of women who were admitted in obstetrical/gynecology department for delivery or obstetrical complications between February 2014 and February 2020. We also studied the impact of the TSAM intervention on health outcomes of sick newborns who were admitted in neonatology unit (neonatal admissions) and live newborns (within 30 minutes of birth) in delivery of Rwandan district hospitals from February 2014 to February 2020. Our unit of observation is the hospital-month. Our final study cohort included 10 intervention hospitals and 15 concurrent control hospitals that did not have ongoing clinical mentorship and / or quality improvement interventions for MNH implemented around the start of the TSAM clinical mentorship (Appendix Figure 1)

### 2.5. Measures

As the TSAM clinical mentorship activities focused on improving maternal and newborn health outcomes, we examined the impact of the TSAM intervention on several measures corresponding to maternal and neonatal mortality and morbidity subdomains of outcomes of care or quality impacts:

- Mortality measures: obstetrical complication case fatality rate, stillbirth rate, 30-minute neonatal mortality rate, and neonatal mortality rate.
- Morbidity measures: Incidence of postpartum hemorrhage (PPH) and neonatal asphyxia.

We also evaluated the impact of the TSAM intervention on other measures:

- Referral rate: Number of referrals to higher level of care per 1000 hospital admissions.
- Caesarean delivery rate: Number of caesarean deliveries per 100 hospital deliveries.

It should be noted that referral and caesarean delivery were not the target of the TSAM clinical mentorship intervention; however, we would argue that improvement in quality of care as a result of the TSAM clinical mentorship intervention could impact these measures. It was not clear a priori which direction referrals would move as a result of quality improvement, and whether the impact would be gradual or immediate. As for caesarean delivery, the direction of the effect would be dependent on whether it was previously underused or overused. In the former scenario, the intervention might increase the caesarean delivery rate, whereas in the latter, it might decrease it.

### 2.6. Design and Analytical strategy

We employed controlled interrupted time series (CITS) design^38, 39^ to examine the impact of the TSAM intervention on outcome measures described above. This approach assumes that the timing of the intervention implementation was quasi-random, i.e. it did not coincide with other events driving the outcomes described above, except salient temporal confounding concerns occurring in both TSAM intervention and concurrent control hospitals. As such, we assumed that absent the TSAM intervention the rates in the outcome measures in the intervention group would have changed in the same way as the change in the concurrent control group.^40^

Given the TSAM clinical mentorship intervention started across hospitals at two different times in 2017, we used study time (the number of months relative to each hospital’s month of the TSAM clinical mentorship start) instead of calendar time, allowing each hospital to be standardized to the others based on their individual index month. In this CITS, we used location-based controls, which consisted of hospitals in which TSAM was not implemented.^41^ Each control hospital was assigned one of the index months (June or November 2017) randomly.^40^ To fit our statistical models, we first calculated the mortality, caesarean delivery, and referral rates across all hospital-months (or hospital-quarters for obstetrical complication case fatality rate because maternal death is a relatively rare event making monthly estimates too noisy) in our study cohort. We then used these aggregate figures to fit segmented linear regression models to estimate immediate (or level) and over time (trend) changes in rates of each of our outcome measures. We provided details about our statistical models in Appendix (see Appendix page 1). Segmented regression models for mortality measures were adjusted for referrals to higher level of care.

Notably, the stillbirth case definition in Rwanda changed from gestation age (GA) > 22 weeks for the period before 2018 to GA ≥ 28 weeks from 2018 and onward. This change in the stillbirth case definition coincided with the TSAM implementation period, affecting the level and / or trend of times series of stillbirth rate in both groups. As the trends of stillbirth rate in intervention and control hospitals did not appear linear during the implementation of TSAM, we calculated the ratio of, and difference in, stillbirth rate between intervention and control hospitals. We then used segmented linear regression model of single interrupted time series (SITS) to estimate the impact of the TSAM intervention on ratios and differences in (intrapartum) stillbirth rate between intervention and control hospitals. This approach has previously been used in other research.^42^

As successive observations may have been correlated over time, we assessed for autocorrelation using the Durbin-Watson test and visual plots (autocorrelation function and partial autocorrelation function) and controlled for autocorrelation using appropriate adjustments in a generalized least squares model.^38^ Moreover, we used likelihood ratio tests to validate the inclusion of autocorrelation structure in each model. We also assessed for seasonality in the outcome measures and, where significant, appropriate adjustments by deseasonalizing data prior to analysis, which was achieved by decomposing the time series data into trend, seasonal, and noise components, and then subtracting the seasonal component.^43^ Similarly, we checked for non-stationarity using the Augmented Dickey– Fuller test.^44^ Additionally, we identified outliers and treated them as missing observations for each outcome measure when they exceeded five standard deviations from the mean time trend estimated using hospital-level local regression. Missing data were imputed using seasonally decomposed missing value imputation, accounting for seasonal patterns in the time series.^45^ Overall, missing values accounted for less than 5% of observations across our outcome measures. All analyses were performed using R version 4.0.2. ^46^

#### 2.6.1. Subgroup and sensitivity analyses

We performed subgroup analyses, determined a priori, to examine whether the TSAM intervention had similar or differential impact across sub-groups. We had planned to stratify all analyses by North and South; however, given there was too much noise (estimates were very unstable) in resulting time series when stratified this was not pursued. We studied the impact of the TSAM intervention on intrapartum stillbirth rate specifically as the impact of the TSAM intervention on stillbirth rate would have occurred through reduction of intrapartum stillbirth (fresh stillbirth), which is linked to the quality of intrapartum obstetrical care.

We conducted sensitivity analyses to check the robustness of our study results. First, we conducted a CITS with segmented mixed effects models to adjust for clustering of observations from the same hospital and heterogeneity in level and trend across hospitals. Lastly, clinical mentorship such as TSAM that aimed to enhance the uptake of clinical practice guidelines might have affected the existing admission protocols.^47^ As such, we studied longitudinal changes in hospital admissions to assess whether significant change occurred after the start of the TSAM clinical mentorship implementation.

### 2.7. Ethical approval

The study was approved by the University of British Columbia Behavioural Research Ethics Board (Certificate # H18-02591).

### 2.8. Patient and Public Involvement

This research was done without patient involvement. Patients were not invited to comment on the study design and were not consulted to develop patient relevant outcomes or interpret the results. Patients were not invited to contribute to the writing or editing of this document for readability or accuracy.

## 3. Results

Table 1 summarizes the characteristics of our study cohort, which included 25 hospitals, comprising of 339,850 hospital deliveries, 35,932 obstetrical complication related admissions, and 94,584 neonatal admissions (Table 1).

**Table 1.**
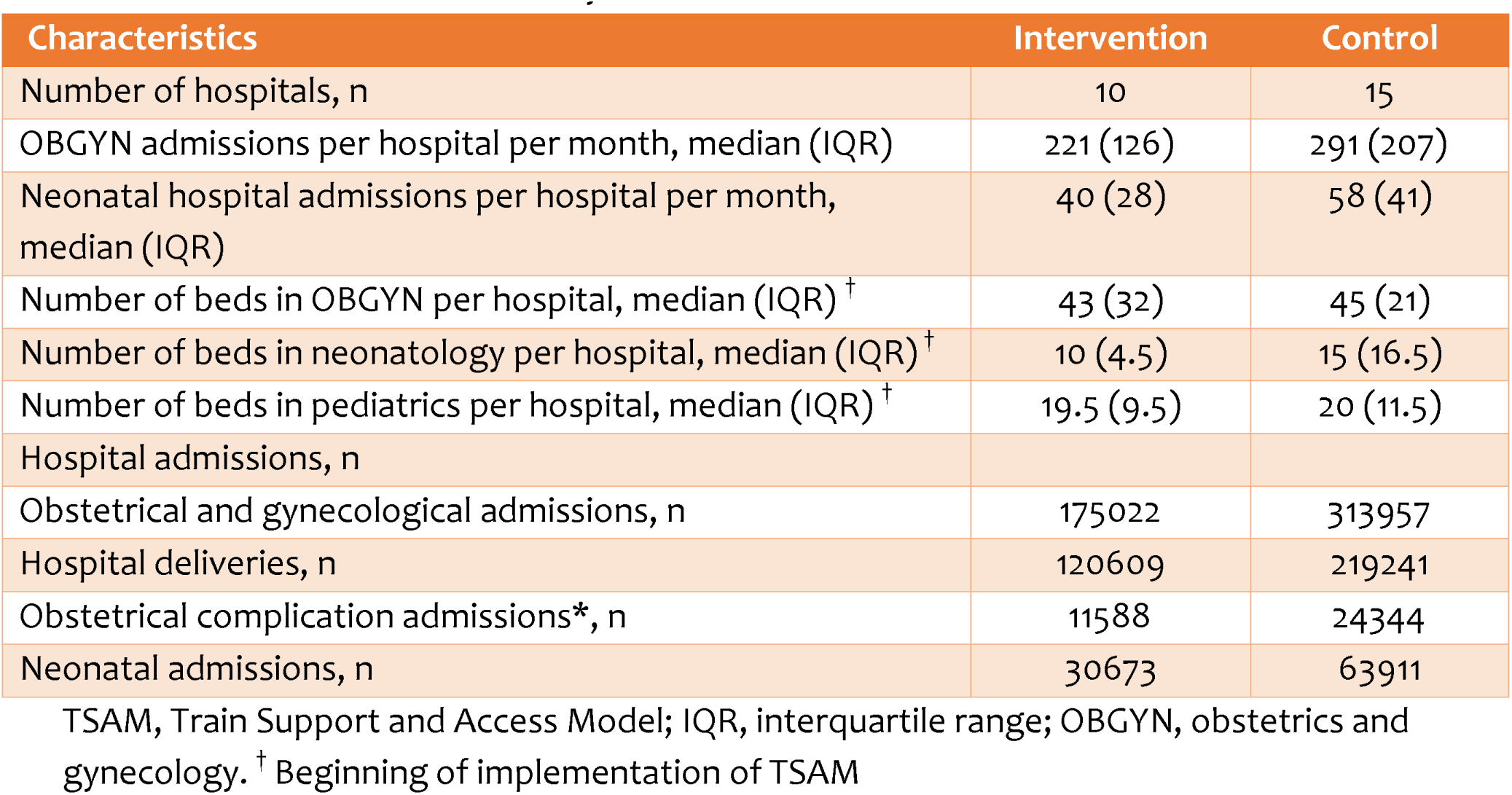
Characteristics of the study cohort.

### 3.1. Impact on hospital mortality

#### 3.1.1. Obstetrical complication case fatality

We found that the implementation of the TSAM intervention had no statistically significant impact on the level of obstetrical complication case fatality rate (level change: –0.03%, 95% CI: –0.73 to 0.66, p=0.92) in intervention hospitals relative to control hospitals (Figure 2 and Table 2). However, the intervention was associated with a significant decrease in the trend of obstetrical complication case fatality rate (trend change: –0.16% / quarter, 95% CI: –0.27 to – 0.05, p=0.007) in intervention hospitals relative to control hospitals (Figure 2 and Table 2). Cumulatively after two years, the implementation of TSAM led to 83.9% fewer obstetrical complication deaths in intervention hospitals than would have been expected absent the intervention.

**Figure 2.**
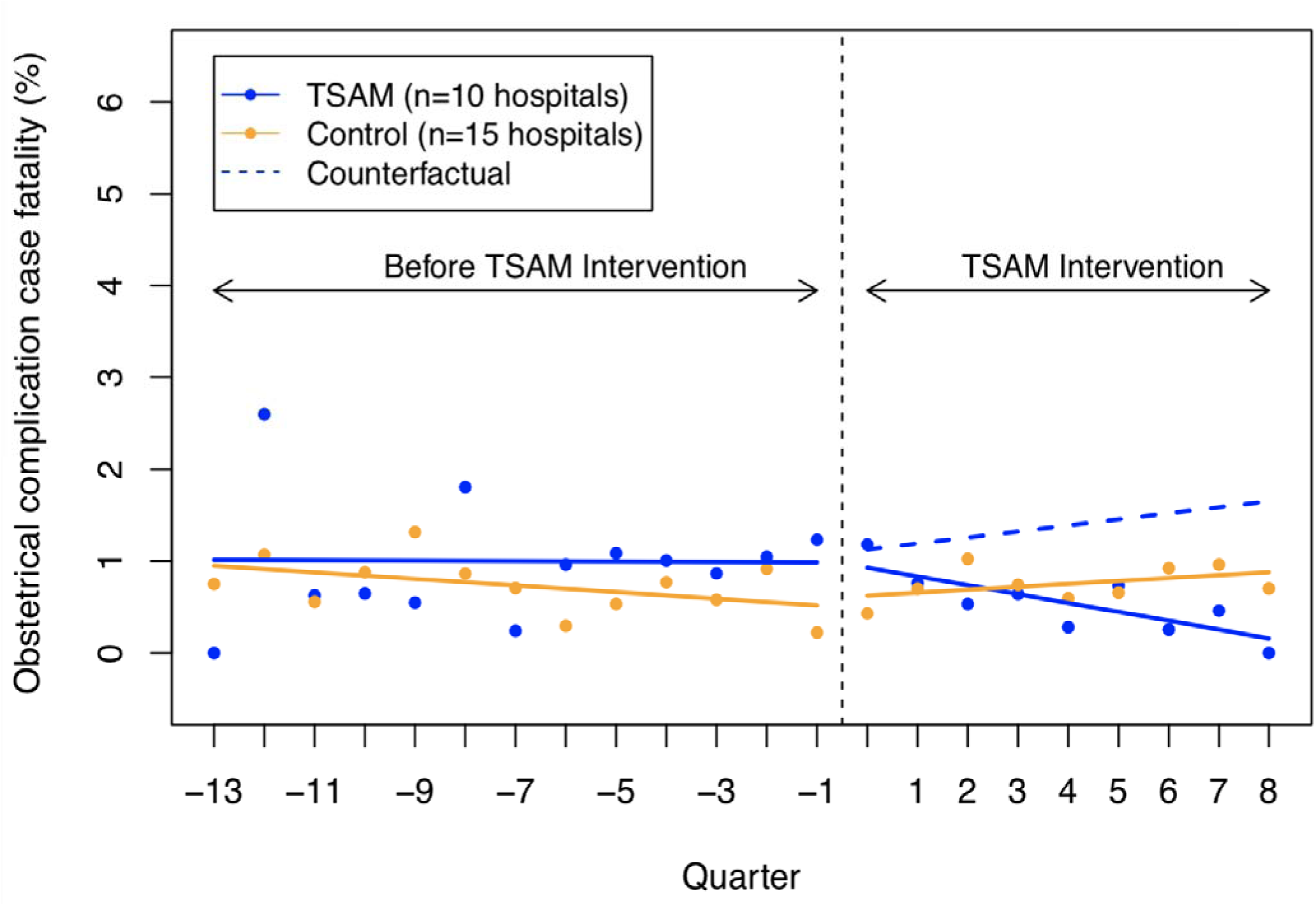
CITS analysis of impact of TSAM on obstetrical complication case fatality rate.

**Table 2.** Results of segmented regression analysis of impact of TSAM on obstetrical complication case fatality and 30-minute neonatal mortality.

|  | Estimates (95% CI) | p value |
| --- | --- | --- |
| **Obstetrical complication case fatality rate (%) \*** |  |  |
| Control, before intervention | 0.98 (0.67, 1.29) | <0.0001 |
| Control's trend, before intervention | -0.03 (-0.07, 0.003) | 0.08 |
| Difference between TSAM and control, before intervention | 0.02 (-0.41, 0.47) | 0.89 |
| Difference in trends between TSAM and control, before intervention | 0.03 (-0.02, 0.08) | 0.24 |
| Control, after intervention | 0.07 (-0.42, 0.56) | 0.77 |
| Control's trend, after intervention | 0.06 (-0.01, 0.14) | 0.10 |
| Difference between TSAM and control after intervention | -0.03 (-0.73, 0.66) | 0.92 |
| Difference in trend between TSAM and control after intervention | -0.16 (-0.27, -0.05) | 0.007 |
| **Neonatal mortality rate /1000 hospital neonatal admissions** |  |  |
| Control, before intervention | 96.35 (85.27, 107.44) | <0.0001 |
| Control's trend, before intervention | -0.49 (-0.86, -0.11) | 0.01 |
| Difference between TSAM and control, before intervention | 2.87 (-9.88, 15.64) | 0.65 |
| Difference in trends between TSAM and control, before intervention | 0.34 (-0.19, 0.87) | 0.21 |
| Control, after intervention | 1.16 (-12.59, 14.93) | 0.86 |
| Control's trend, after intervention | 0.61 (-0.12, 1.35) | 0.10 |
| Difference between TSAM and control after intervention | -8.13 (-27.49, 11.22) | 0.41 |
| Difference in trend between TSAM and control after intervention | -1.10 (-2.15, -0.05) | 0.04 |
| **30-minute neonatal mortality rate / 1000 hospital live births** |  |  |
| Control, before intervention | 7.61 (6.89, 8.34) | <0.0001 |
| Control's trend, before intervention | -0.05 (-0.08, -0.02) | 0.001 |
| Difference between TSAM and control, before intervention | -2.27 (-3.29, -1.24) | <0.0001 |
| Difference in trends between TSAM and control, before intervention | 0.07 (0.03, 0.12) | 0.0007 |
| Control, after intervention | 0.25 (-0.90, 1.41) | 0.66 |
| Control's trend, after intervention | -0.0003 (-0.06, 0.06) | 0.99 |
| Difference between TSAM and control after intervention | -1.24 (-2.88, 0.39) | 0.13 |
| Difference in trend between TSAM and control after intervention | -0.08 (-0.16, 0.006) | 0.07 |

CI, confidence interval; TSAM, training support and access model. <sup>†</sup>\*Estimates were adjusted for referral rate to higher levels of care. \*Hospital admissions due to obstetrical complications include hemorrhage (antepartum hemorrhage and postpartum hemorrhage), (pre)-eclampsia, prolonged labor, abortion, ectopic pregnancy, uterine rupture, anemia in pregnancy, and infections.

#### 3.1.2. Stillbirth rate

As described earlier, the change in stillbirth case definition during the TSAM implementation period affected the level and / or trend of times series of stillbirth rate in both groups as can be seen in Figure 3. We found that the implementation of TSAM had no statistically significant impact on the level or trend of ratio of stillbirth incidence rate between intervention and control hospitals rate ratio (level change: 0.02, 95% CI: –0.11 to 0.15, p=0.75; trend change: –0.001 / month, 95% CI: –0.009 to 0.005, p=0.62) (Appendix Figure 2 and Appendix Table 1). Similarly, the TSAM intervention had no statistically significant impact on the level or trend of difference in stillbirth rate between intervention and control hospitals (level change: 2.04 stillbirths / 1000 births, 95% CI: –2.97 to 7.07, p=0.41; trend change: –0.02 stillbirths / 1000 births / month, 95% CI: –0.29 to 0.25, p=0.86) (Appendix Figure 3 and Appendix Table 1).

**Figure 3.**
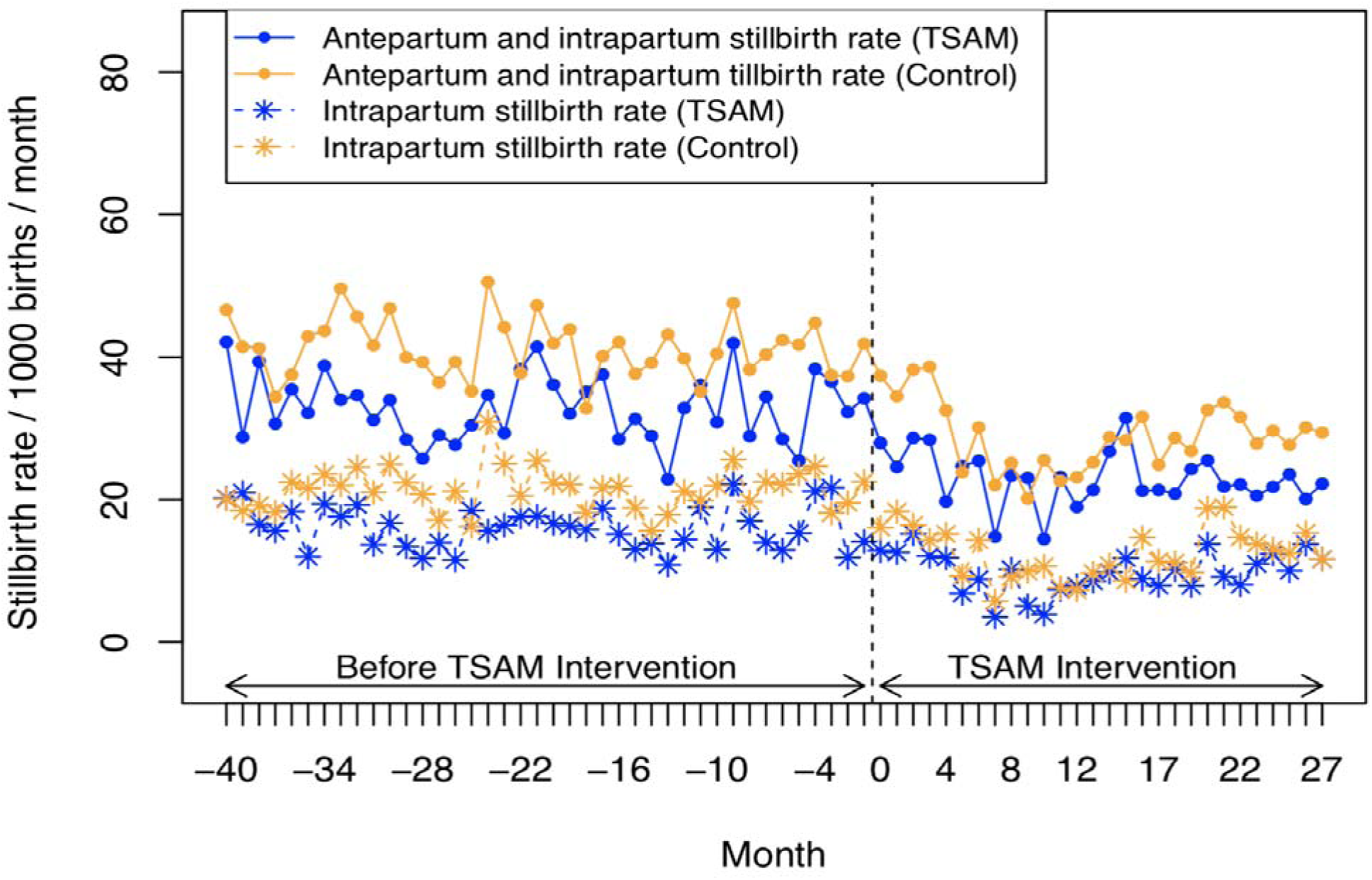
Times series of monthly stillbirth rate.

We also examined the impact of the TSAM intervention specifically on ratio and difference in intrapartum stillbirth rate between intervention and control hospitals. The implementation of TSAM had no statistically significant impact on the level or trend of ratio of intrapartum stillbirth rate between intervention and control hospitals (level change: 0.04, 95% CI: –0.14 to 0.23, p=0.63; trend change: 0.004 / month, 95% CI: –0.006 to 0.014, p=0.41) (Appendix Figure 4 and Appendix Table 1). Similarly, the TSAM intervention had no statistically significant impact on the level or trend of difference in intrapartum stillbirth rate between intervention and control hospitals (level change: 3.18 stillbirths / 1000 births, 95% CI: –0.28 to 6.65, p=0.07; trend change: 0.04 stillbirths / 1000 births / month, 95% CI: –0.14 to 0.23, p=0.61) (Appendix Figure 5 and Appendix Table 1).

**Figure 4.**
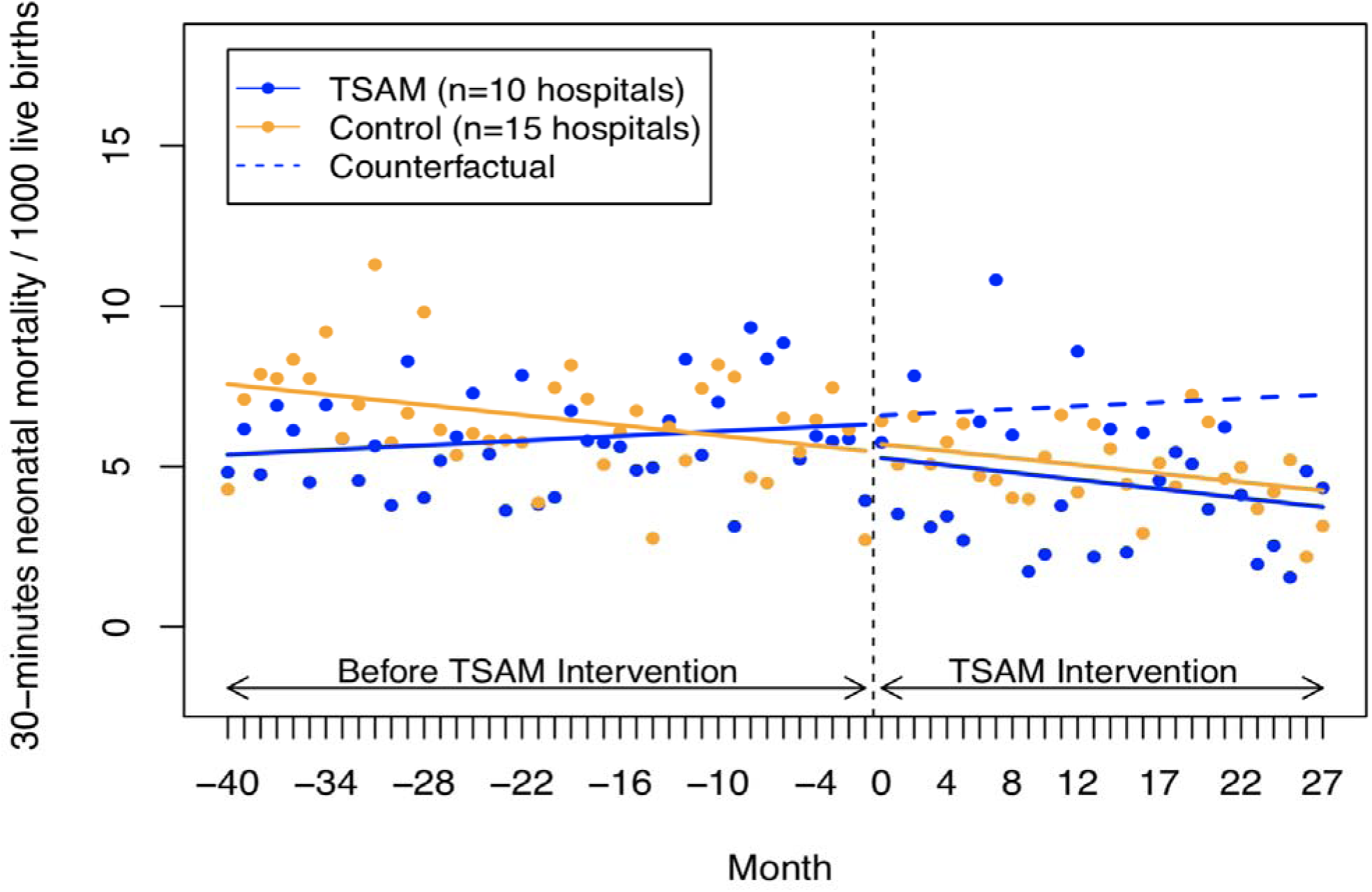
CITS analysis of impact of TSAM intervention on 30-minutes neonatal mortality.

**Figure 5.**
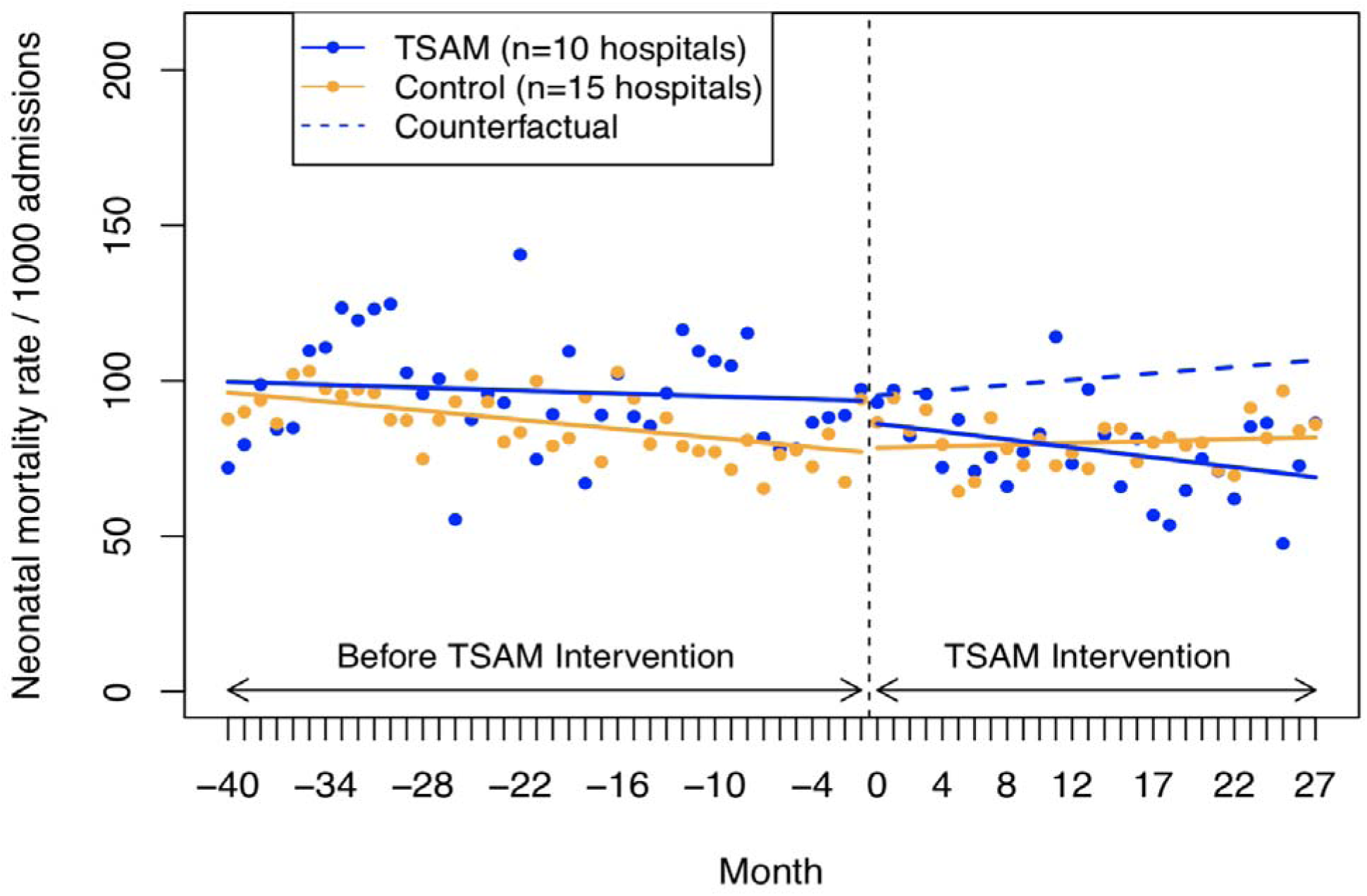
CITS analysis of impact of TSAM intervention on neonatal mortality rate.

#### 3.1.3. Neonatal mortality

We found that the implementation of the TSAM intervention was associated with an immediate decrease in the neonatal mortality rate (level change: –8.13 deaths / 1000 admissions, 95% CI: –27.49 to 11.22, p=0.41), although not statistically significant, in intervention hospitals relative to control hospitals (Figure 5 and Table 2). Similarly, the TSAM intervention was associated with a significant reduction in the trend of neonatal mortality rate (trend change: –1.10 deaths / 1000 admissions / month, 95% CI: –2.15 to –0.05, p=0.04) in intervention hospitals compared with control hospitals (Figure 5 and Table 2). Cumulatively after two years, the implementation of TSAM led to 32.1 % fewer neonatal deaths in intervention hospitals than would have been expected absent the intervention.

Looking specifically at the 30-minute neonatal mortality rate, we found that the intervention was associated with a decrease in the level and trend of rate of newborn death within 30 minutes of birth (level change: –1.24 newborn deaths / 1000 live births, 95% CI: –2.88 to 0.39, p=13; trend change: –0.08 newborn deaths / 1000 live births / month, 95% CI: –0.16 to 0.006, p=0.07), although not statistically significant, in intervention hospitals compared with control hospitals (Figure 4 and Table 2). Cumulatively after two years, the implementation of TSAM led to 44.5% fewer newborn deaths within 30 minutes of birth in intervention hospitals than would have been expected absent the intervention.

### 3.2. Impact on hospital morbidity

Our morbidity indicators included incidence of postpartum hemorrhage (PPH) and neonatal asphyxia. We found that the TSAM intervention was associated with a significant decrease in the trend of PPH incidence rate (trend change: –0.40 PPH cases / 1000 deliveries / month, 95% CI: –0.70 to –0.09, p=0.01) in intervention hospitals relative to control hospitals (Figure 6A and Table 3). Cumulatively after two years, the implementation of TSAM led to 30.3% fewer PPH cases in intervention hospitals than would have been expected absent the intervention. Similarly, our results showed that the implementation of TSAM was associated with a significant decrease in the level and trend of neonatal asphyxia incidence rate (level change: –10.80 neonatal asphyxia / 1000 live births, 95% CI: –21.61 to 0.006, p=0.05; trend change: –1.10 neonatal asphyxia / 1000 live births / month, 95% CI: –1.70 to –0.49, p=0.0005) (Figure 6B and Table 3). Cumulatively after two years, the implementation of TSAM led to 47.9% fewer neonatal asphyxia cases in intervention hospitals than would have been expected absent the intervention.

**Figure 6.**
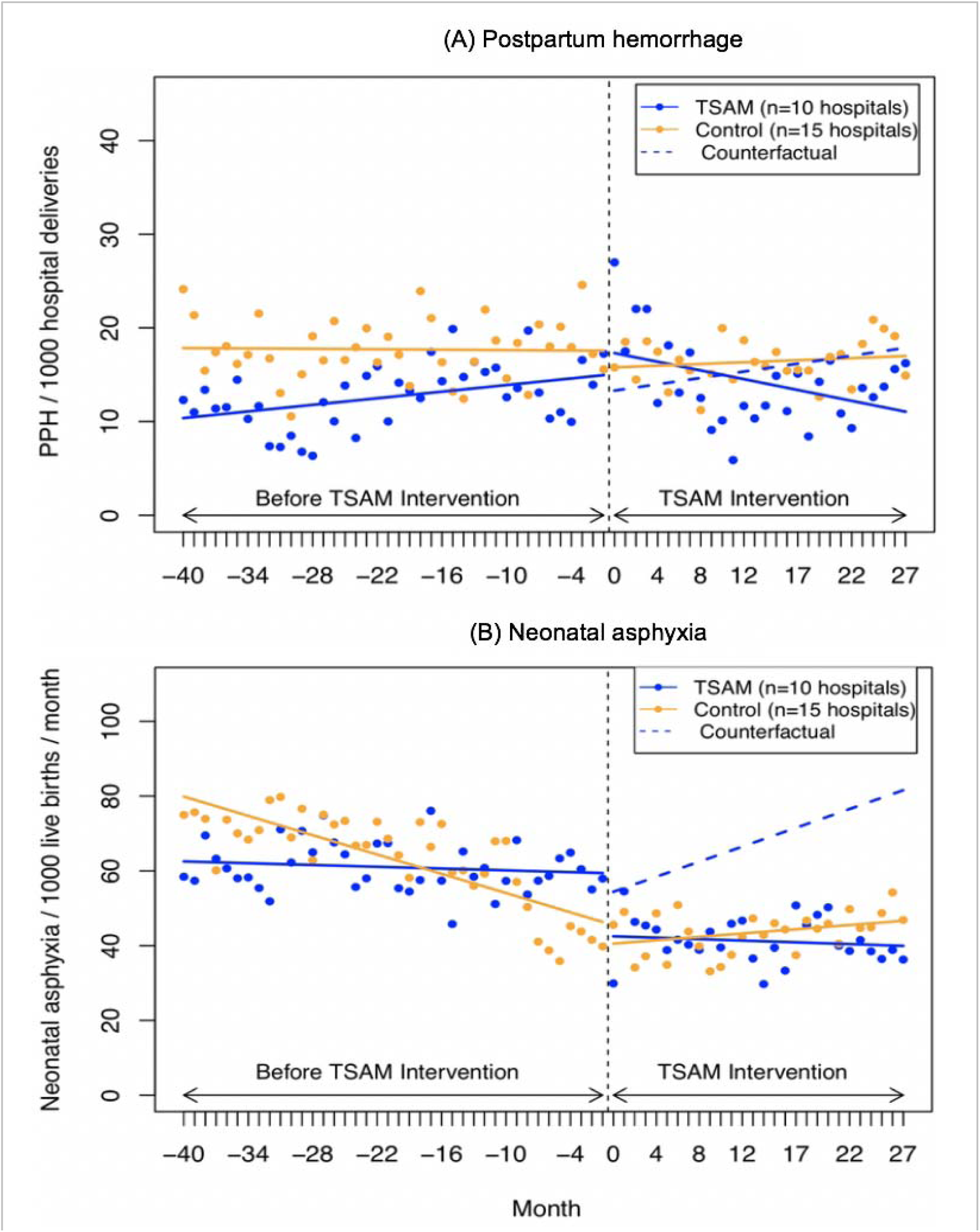
CITS analysis of impact of TSAM intervention on PPH incidence rate (A) neonatal asphyxia incidence rate (B).

**Table 3.** Results of segmented regression analysis of impact of SAM on postpartum hemorrhage and neonatal asphyxia incidence rates.

|  | Estimates (95% CI) | p value |
| --- | --- | --- |
| **Postpartum hemorrhage incidence rate / 1000 deliveries** |  |  |
| Control, before intervention | 17.85 (15.27, 20.43) | <0.0001 |
| Control's trend, before intervention | -0.007 (-0.11, 0.10) | 0.89 |
| Difference between TSAM and control, before intervention | -7.60 (-11.25, -3.95) | 0.0001 |
| Difference in trends between TSAM and control, before intervention | 0.12 (-0.02, 0.27) | 0.11 |
| Control, after intervention | -1.82 (-5.68, 2.03) | 0.35 |
| Control's trend, after intervention | 0.05 (-0.16, 0.26) | 0.64 |
| Difference between TSAM and control after intervention | 4.40 (-1.05, 9.87) | 0.11 |
| Difference in trend between TSAM and control after intervention | -0.40 (-0.70, -0.09) | 0.01 |
| **Neonatal asphyxia incidence rate / 1000 live births** |  |  |
| Control, before intervention | 80.71 (75.61, 85.81) | <0.0001 |
| Control's trend, before intervention | -0.85 (-1.07, -0.64) | <0.0001 |
| Difference between TSAM and control, before intervention | -18.10 (-25.31, -10.89) | <0.0001 |
| Difference in trends between TSAM and control, before intervention | 0.77 (0.47, 1.08) | <0.0001 |
| Control, after intervention | -6.02 (-13.67, 1.61) | 0.12 |
| Control's trend, after intervention | 1.08 (0.65, 1.51) | <0.0001 |
| Difference between TSAM and control after intervention | -10.80 (-21.61, 0.006) | 0.05 |
| Difference in trend between TSAM and control after intervention | -1.10 (-1.70, 0.49) | 0.0005 |

CI, confidence interval; TSAM, training support and access model

### 3.3. Referral and Caesarian delivery

We found that the implementation of the TSAM clinical mentorship had no statistically significant impact on the level or trend of maternal referral rate, neonatal, and pediatric referral rate in intervention hospitals relative to control hospitals (Appendix Figures 6-8 and Appendix Table 2). However, the intervention was associated with a significant increase in the trend of caesarean delivery rate (trend change: 0.21% / month, 95% CI: 0.07 to 0.35, p=0.002) in intervention hospitals relative to control hospitals (Appendix Figure 9 and Appendix Table 3). Cumulatively after two years, the implementation of TSAM led to 11.3% more caesarean deliveries in intervention hospitals than would have been expected absent the intervention.

### 3.4. Sensitivity analyses

All the additional analyses showed substantively similar results. Implementation of the TSAM intervention was not associated with significant changes in the level or trend of hospital admission rate (i.e., obstetrical and gynecology admissions and neonatal and pediatric admissions) and hospital delivery rate in intervention hospitals relative to control hospitals, assuaging concerns about selection-instrumental threat due to significant change in intervention hospital admission rate and or delivery rate as a result of the intervention (Appendices Figures 10-12 and Appendix Table 4).

## 4. Discussion

The TSAM clinical mentorship was implemented in ten Rwandan district hospitals in an effort to improve outcomes of care for mothers and newborns. Using a rigorous quasi-experimental interrupted time series design and routine health information system data, we evaluated the impact of the TSAM clinical mentorship intervention. We found that the implementation of the TSAM clinical mentorship intervention was associated with a reduction in the 30-minute neonatal mortality rate, neonatal mortality rate, and obstetrical complication case fatality rate in intervention hospitals relative to control hospitals. Similarly, TSAM was associated with a reduction in the incidence of postpartum hemorrhage and neonatal asphyxia. However, the (intrapartum) stillbirth rate did not decline as a result of the TSAM intervention.

While multiple studies have evaluated the impact of mentorship or coaching on processes of care quality, there is a dearth of rigorous evidence on the impact of these interventions on outcomes of care for mothers and newborns. The BetterBirth trial conducted in India assessed the impact of an 8-month coaching-based WHO safe childbirth checklist program on processes and outcomes of care for mothers and newborns in primary care facilities.^13^ Consistent with our evaluation of the TSAM clinical mentorship program, they found that the program did not reduce the stillbirth rate. However, the coaching-based WHO safe childbirth checklist program improved processes of care quality including adherence to essential birth practices, but not sufficiently enough to improve outcomes of care and the level of adherence decreased over time after coaching stopped.^13^ A recent systematic review found that mentoring programs targeting improvements in health providers’ competence and institutional performance in Africa were associated with improved processes of care for infectious diseases, and maternal, neonatal, and childhood illnesses.^12^ Similarly, studies that evaluated the impact of mentorship in Rwanda, although they have focused on primary-level health centers, have found that mentorship for nurses is associated with improved processes of care quality for MNCH conditions and for NCDs such as diabetes and mental health conditions.^48–50^ Even though we could not implicitly study the impact of the TSAM clinical mentorship on processes of care meaningfully because many relevant processes of care quality indicators were not available in the Rwanda HMIS when the intervention was implemented, we would argue that TSAM improved maternal and newborn health outcomes through improvements in processes of care quality.

Unlike the coaching-based WHO safe childbirth checklist program that ceased coaching after eight months, the TSAM clinical mentorship had been ongoing since its initiation until the start of the COVID-19 outbreak in the country in March 2020 and not only were better practices supported and MNCH emergency cases reviewed by both mentees and mentors together, but health providers were also mentored to enhance their clinical skills to provide quality care. Mentee performance and progress were tracked using logbooks. Moreover, the TSAM clinical mentorship focused on improving clinical quality beyond only adhering to the tasks outlined in the WHO safe childbirth checklist. Additionally, unlike the BetterBirth study that was done in primary care facilities—which generally lack capacity to provide advance care should complications arise during childbirth—, TSAM was implemented in hospitals that can provide both basic care and care for complications arising during childbirth. Further, TSAM involved the leadership of hospitals in the continuous quality improvement to facilitate successful implementation. All these may explain why the TSAM clinical mentorship program had a positive impact on mortality and morbidity indicators.

Our analyses showed that the TSAM clinical mentorship was associated with an increase in the trend of caesarean delivery rate. This increase along with improvements in maternal and newborn health outcomes suggest that the caesarean delivery was likely underused previously. Whereas the incidence of neonatal asphyxia, 30-minute neonatal mortality, and neonatal mortality decreased as a result of TSAM, the incidence of intrapartum stillbirth rate did not change. It is not clear why the incidence of intrapartum stillbirth rate did not decrease. However, a potential explanation for this could be that intrapartum stillbirth rate did not decrease because of inherent delays in referrals of women in labor from lower levels of care to the hospitals. Additionally, it could be that the quality of intrapartum care did improve but not significant enough to reduce the incidence of intrapartum stillbirth rate at the TSAM hospitals. Clearly, future research will be required to shed more light on these explanations to help us understand why the intrapartum stillbirth rate did not decline and potentially inform improvement to the TSAM clinical mentorship as it is scaled up.

## Strengths and limitations

While a well-designed randomized controlled trial would arguably provide the strongest evidence on the impact of the TSAM clinical mentorship intervention, it was not possible to randomize this intervention as it had already been implemented. Given this, the best alternative option to evaluate the impact of this intervention was to use a rigorous quasi-experimental interrupted time series design with a concurrent control group.^39^ Indeed, potential changes in health services utilization, hospital resources, and the health status of admitted mothers, newborns and children would bias my findings only if they coincidentally changed when the implementation of the TSAM intervention was started and also differentially between intervention and control. Sensitivity analyses showed that implementing the TSAM clinical mentorship generally had no statistically significant impact on the level or trend of hospital admission rates. Similarly, other clinical mentorship programs for maternal, newborn, and child health could have biased our findings if their implementation coincided with the implementation of the TSAM clinical mentorship program. However, we are not aware of any programs that were being implemented during TSAM. Indeed, the suddenness of changes generally observed at the time of the TSAM clinical mentorship intervention make it less plausible that the changes were caused by factors other than TSAM or that it was a random fluctuation over time. Moreover, our analyses accounted for all key methodological considerations (e.g., autocorrelation) that could have biased estimates. Further, there are other relevant indicators (e.g., morbidity indicators such as perineal tear and fistula, and processes of care quality indicators) that we could have examined were not captured in the Rwanda HMIS. Lastly, the change in the case definition of stillbirths during the study period may have introduced misclassification bias.

## Conclusion

In summary, using a rigorous quasi-experimental design and routine health information system data, we showed that a quality improvement strategy that employed continuous quality improvement approaches using onsite clinical mentorship of health providers along with involvement of health facility leadership to facilitate the improvement was associated with improvements in maternal and newborn health outcomes in Rwanda. Our findings provide evidence that can inform the scale up of TSAM across the country and other settings within similar contexts to contribute to improving maternal and newborn health outcomes in the SDG era and beyond. However, there still is a need for specific interventions to reduce stillbirth rate in Rwanda. Moreover, given it is critical that low-income countries like Rwanda, with scarce resources, allocate available resources toward scaling up the most cost-effective interventions, studies evaluating the cost-effectiveness of the TSAM clinical mentorship intervention will complement the present analysis and inform its scale up. Similarly, given the importance of context and the reality that interventions are rarely implemented with perfect fidelity under real-world conditions, it is critical to regularly track the real-world experiences with the implementation of TSAM in order to identify the potential barriers and facilitators to its successful implementation including adoption and scale-up over time.

## Supporting information

TSAM supplemental file

## Data Availability

All data produced in the present work are contained in the manuscript.

## Notes

### Competing Interest Statement

Drs. Hategeka, Lynd, Kenyon, Ngabonzima, Luginaah and Cechetto were involved in the development and / or implementation of TSAM in Rwanda.

### Funding Statement

Dr. Hategeka received support through a Vanier Canada Graduate Scholarship and a Banting Postdoctoral Fellowship from the Canadian Institutes of Health Research. Dr. Cechetto was awarded the grant from Global Affairs Canada that supported the development and implementation of TSAM in Rwanda. Dr. Law received salary support through a Canada Research Chair in Access to Medicines and a Michael Smith Foundation for Health Research Scholar Award.

### Author Declarations

The University of British Columbia Behavioural Research Ethics Board gave ethical approval for this work (Certificate # H18-02591).

