## Supplementary material for "Impact of the training, support and access model (TSAM) on patient health outcomes in Rwanda: controlled longitudinal study": TSAM supplemental file: Hategeka et al TSAM Paper Supplementary Appendix.docx

**Statistical appendix**

The model structure had the following general form to model each outcome measure for the TSAM intervention status *j*, at time *t*:

$${Outcome}_{jt}=\beta_{0}+\beta_{1}\cdot{time}_{t}+\beta_{2}\cdot{TSAM}_{j}+\beta_{3}\cdot{TSAM}_{j}\cdot{time}_{t}+\beta_{4}\cdot{level}_{t}+\beta_{5}\cdot{trend}_{t}+\beta_{6}\cdot{level}_{t}\cdot{TSAM}_{j}+\beta_{7}\cdot{trend}_{t}\cdot{TSAM}_{j}+\varepsilon_{jt}$$

where *outcome* represents each of the outcome measures described above; *time* is the month (or quarter) in study time *t* (i.e. 1, 2, 3, 4…); *TSAM* is an indicator for whether the hospital had implemented the TSAM intervention at time *t*; *level* represents whether the TSAM intervention was in place in month (or quarter) *t*; and *trend* represents the month (or quarter) *t* it was after the hospital had implemented the TSAM intervention. In each CITS model, *β_0_* estimates the baseline level of each outcome measure for the control; *β_1_* estimates pre-intervention trend for the control; *β_2_* estimates the baseline difference between control and intervention; *β_3_* estimates the pre-existing difference in trend between intervention and control; *β_4_* and *β_5_* estimate the level and trend changes for the control post-intervention respectively. The two coefficients of interest are *β_6_*, which indicates any immediate change in the *level* of the outcome in the intervention relative to the control, and *β_7_*, which indicates any change in the *trend* of the outcome in the intervention relative to the control following implementation of the intervention. Statistically significant values for these coefficients (*β_6_* and/or *β_7_*) would indicate that the intervention had an impact on the outcome measures evaluated. *ε_jt_* is the error term representing the variability not explained by the model. Using established approaches, we estimated relative changes in the outcomes, wherever significant, in order to make the results easily interpretable.^51^

Appendix Table 1. Results of segmented regression analysis of impact of TSAM on stillbirth

|  | Estimates (95% CI) | *p* value |
| --- | --- | --- |
| Ratio of stillbirth rate, TSAM and control |  |  |
| Baseline level (intercept) | 0.80 (0.71, 0.88) | <0.0001 |
| Trend, before intervention | 0.00014 (-0.003, 0.003) | 0.93 |
| Level change | 0.02 (-0.11, 0.15) | 0.75 |
| Trend change | -0.001 (-0.009, 0.005) | 0.62 |
| Difference in stillbirth rate, TSAM and control |  |  |
| Baseline level (intercept) | -8.59 (-11.84, -5.33) | <0.0001 |
| Trend, before intervention | 0.01 (-0.12, 0.15) | 0.82 |
| Level change | 2.04 (-2.97, 7.07) | 0.41 |
| Trend change | -0.02 (-0.29, 0.25) | 0.86 |
| Ratio of intrapartum stillbirth rate, TSAM and control |  |  |
| Baseline Level (intercept) | 0.80 (0.68, 0.92) | <0.0001 |
| Trend, before intervention | -0.001 (-0.007, 0.003) | 0.45 |
| Level change | 0.04 (-0.14, 0.23) | 0.63 |
| Trend change | 0.004 (-0.006, 0.01) | 0.41 |
| Difference in intrapartum stillbirth rate, TSAM and control |  |  |
| Baseline level (intercept) | -4.51 (-6.76, -2.26) | 0.0002 |
| Trend, before intervention | -0.03 (-0.13, 0.05) | 0.42 |
| Level change | 3.18 (-0.28, 6.65) | 0.07 |
| Trend change | 0.04 (-0.14, 0.23) | 0.61 |

CI, confidence interval; TSAM, training support and access model.

Appendix Table 2. Results of segmented regression analysis of impact of TSAM on referral rate to higher levels for emergency care for newborns with complications at birth and women with obstetrical complications

|  | **Estimates (95% CI)** | ***p* value** |
| --- | --- | --- |
| **Maternal referral rate / 100 obstetrical complication admissions** |  |  |
| Control, before intervention (*β_0_*) | 13.35 (9.45, 17.25) | <0.0001 |
| Control’s trend, before intervention (*β_1_*) | 0.18 (0.02, 0.35) | 0.02 |
| Difference between TSAM and control, before intervention (*β_2_*) | 0.07 (-5.44, 5.58) | 0.97 |
| Difference in trends between TSAM and control, before intervention (*β_3_*) | 0.07 (-0.16, 0.30) | 0.54 |
| Control, after intervention (*β_4_*) | -0.49 (-6.30, 5.32) | 0.86 |
| Control’s trend, after intervention (*β_5_*) | 0.13 (-0.19, 0.46) | 0.42 |
| Difference between TSAM and control after intervention (*β_6_*) | 4.89 (-3.32, 13.11) | 0.24 |
| Difference in trend between TSAM and control after intervention (*β_7_*) | -0.28 (-0.75, 0.17) | 0.22 |
| **Referral rate of newborns with complications / 1000 live births** |  |  |
| Control, before intervention (*β_0_*) | 11.10 (6.32, 15.87) | <0.0001 |
| Control’s trend, before intervention (*β_1_*) | -0.22 (-0.42, -0.02) | 0.02 |
| Difference between TSAM and control, before intervention (*β_2_*) | 1.28 (-5.46, 8.03) | 0.70 |
| Difference in trends between TSAM and control, before intervention (*β_3_*) | 0.18 (-0.09, 0.47) | 0.19 |
| Control, after intervention (*β_4_*) | 5.05 (-1.87, 11.97) | 0.15 |
| Control’s trend, after intervention (*β_5_*) | 0.36 (-0.03, 0.77) | 0.07 |
| Difference between TSAM and control after intervention (*β_6_*) | -2.67 (-12.46, 7.12) | 0.59 |
| Difference in trend between TSAM and control after intervention (*β_7_*) | -0.39 (-0.96, 0.16) | 0.17 |
| **Neonatal referral rate** / 1000 neonatal hospital admissions |  |  |
| Control, before intervention (*β_0_*) | 23.26 (19.32, 27.19) | <0.0001 |
| Control’s trend, before intervention (*β_1_*) | -0.19 (-0.36, -0.02) | 0.02 |
| Difference between TSAM and control, before intervention (*β_2_*) | 11.03 (5.47, 16.59) | 0.0002 |
| Difference in trends between TSAM and control, before intervention (*β_3_*) | -0.18 (-0.42, 0.04) | 0.12 |
| Control, after intervention (*β_4_*) | 5.98 (-0.04, 11.91) | 0.05 |
| Control’s trend, after intervention (*β_5_*) | 0.03 (-0.29, 0.36) | 0.84 |
| Difference between TSAM and control after intervention (*β_6_*) | 4.05 (-4.34, 12.44) | 0.34 |
| Difference in trend between TSAM and control after intervention (*β_7_*) | 0.16 (-0.30, 0.62) | 0.49 |

CI, confidence interval; TSAM, training support and access model.

Appendix Table 3. Results of segmented regression analysis of impact of TSAM on rates of cesarean delivery

|  | **Estimates (95% CI)** | ***p* value** |
| --- | --- | --- |
| **Caesarean delivery rate** / 100 hospital deliveries |  |  |
| Control, before intervention (*β_0_*) | 37.83 (36.67, 39.00) | <0.0001 |
| Control’s trend, before intervention (*β_1_*) | 0.006 (-0.04, 0.05) | 0.80 |
| Difference between TSAM and control, before intervention (*β_2_*) | 7.18 (5.54, 8.83) | <0.0001 |
| Difference in trends between TSAM and control, before intervention (*β_3_*) | -0.14 (-0.21, -0.07) | 0.0001 |
| Control, after intervention (*β_4_*) | 1.55 (-0.29, 3.40) | 0.10 |
| Control’s trend, after intervention (*β_5_*) | 0.10 (0.004, 0.20) | 0.04 |
| Difference between TSAM and control after intervention (*β_6_*) | -0.71 (-3.33, 1.90) | 0.59 |
| Difference in trend between TSAM and control after intervention (*β_7_*) | 0.21 (0.07, 0.35) | 0.002 |

CI, confidence interval; TSAM, training support and access model.

Appendix Table 4. Results of segmented regression analysis of impact of TSAM on hospital admission rate

|  | **Estimates (95% CI)** | ***p* value** |
| --- | --- | --- |
| **Hospital delivery rate / 100 deliveries** |  |  |
| Control, before intervention (*β_0_*) | 29.76 (28.95, 30.57) | <0.0001 |
| Control’s trend, before intervention (*β_1_*) | -0.01 (-0.05, 0.01) | 0.34 |
| Difference between TSAM and control, before intervention (*β_2_*) | 7.86 (6.71, 9.00) | <0.0001 |
| Difference in trends between TSAM and control, before intervention (*β_3_*) | -0.04 (-0.08, 0.007) | 0.09 |
| Control, after intervention (*β_4_*) | 1.17 (-0.004, 2.35) | 0.05 |
| Control’s trend, after intervention (*β_5_*) | 0.15 (0.08, 0.22) | <0.0001 |
| Difference between TSAM and control after intervention (*β_6_*) | 1.04 (-0.62, 2.71) | 0.22 |
| Difference in trend between TSAM and control after intervention (*β_7_*) | 0.004 (-0.09, 0.10) | 0.92 |
| **Neonatal and pediatric admission rate / 1000 population** |  |  |
| Control, before intervention (*β_0_*) | 0.49 (0.42, 0.56) | <0.0001 |
| Control’s trend, before intervention (*β_1_*) | 0.001 (-0.001, 0.004) | 0.34 |
| Difference between TSAM and control, before intervention (*β_2_*) | 0.07 (-0.02, 0.16) | 0.16 |
| Difference in trends between TSAM and control, before intervention (*β_3_*) | 0.001 (-0.002, 0.005) | 0.47 |
| Control, after intervention (*β_4_*) | -0.02 (-0.11, 0.06) | 0.58 |
| Control’s trend, after intervention (*β_5_*) | -0.003 (-0.01, 0.002) | 0.29 |
| Difference between TSAM and control after intervention (*β_6_*) | -0.02 (-0.15, 0.10) | 0.68 |
| Difference in trend between TSAM and control after intervention (*β_7_*) | 0.002 (-0.005, 0.01) | 0.59 |
| **Obstetrical / gynecological admission rate / 1000 population** |  |  |
| Control, before intervention (*β_0_*) | 0.90 (0.86, 0.94) | <0.0001 |
| Control’s trend, before intervention (*β_1_*) | -0.0002 (-0.002, 0.001) | 0.76 |
| Difference between TSAM and control, before intervention (*β_2_*) | 0.25 (0.19, 0.31) | <0.0001 |
| Difference in trends between TSAM and control, before intervention (*β_3_*) | 0.0006 (-0.001, 0.003) | 0.60 |
| Control, after intervention (*β_4_*) | -0.009 (-0.07, 0.05) | 0.77 |
| Control’s trend, after intervention (*β_5_*) | 0.006 (-0.002, 0.009) | 0.0006 |
| Difference between TSAM and control after intervention (*β_6_*) | 0.007 (-0.08, 0.09) | 0.87 |
| Difference in trend between TSAM and control after intervention (*β_7_*) | 0.001 (-0.003, 0.006) | 0.67 |

CI, confidence interval; TSAM, training support and access model.

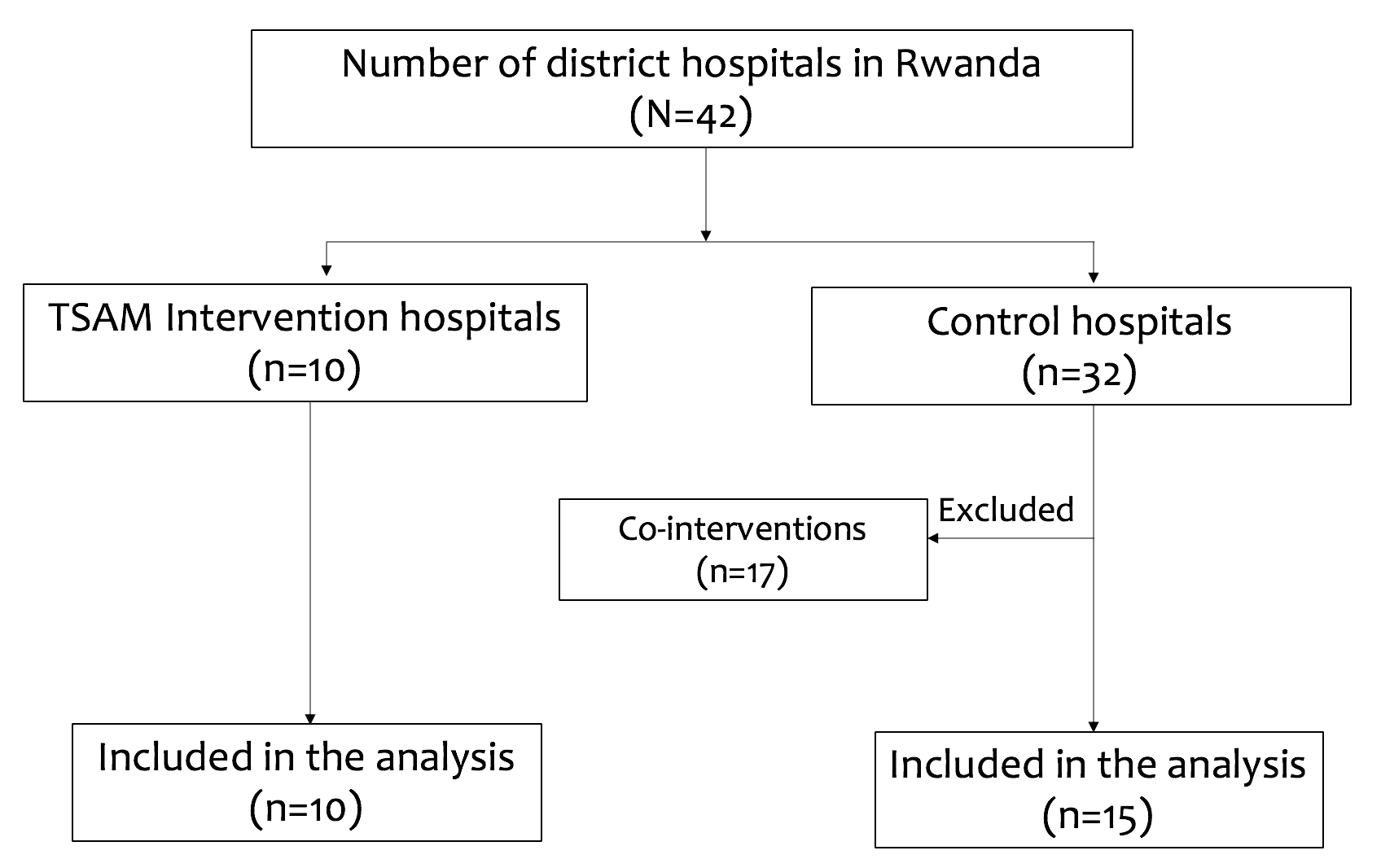

Appendix Figure 1. Flowchart for selection of the study cohort, impact of TSAM

TSAM, training support and access model.

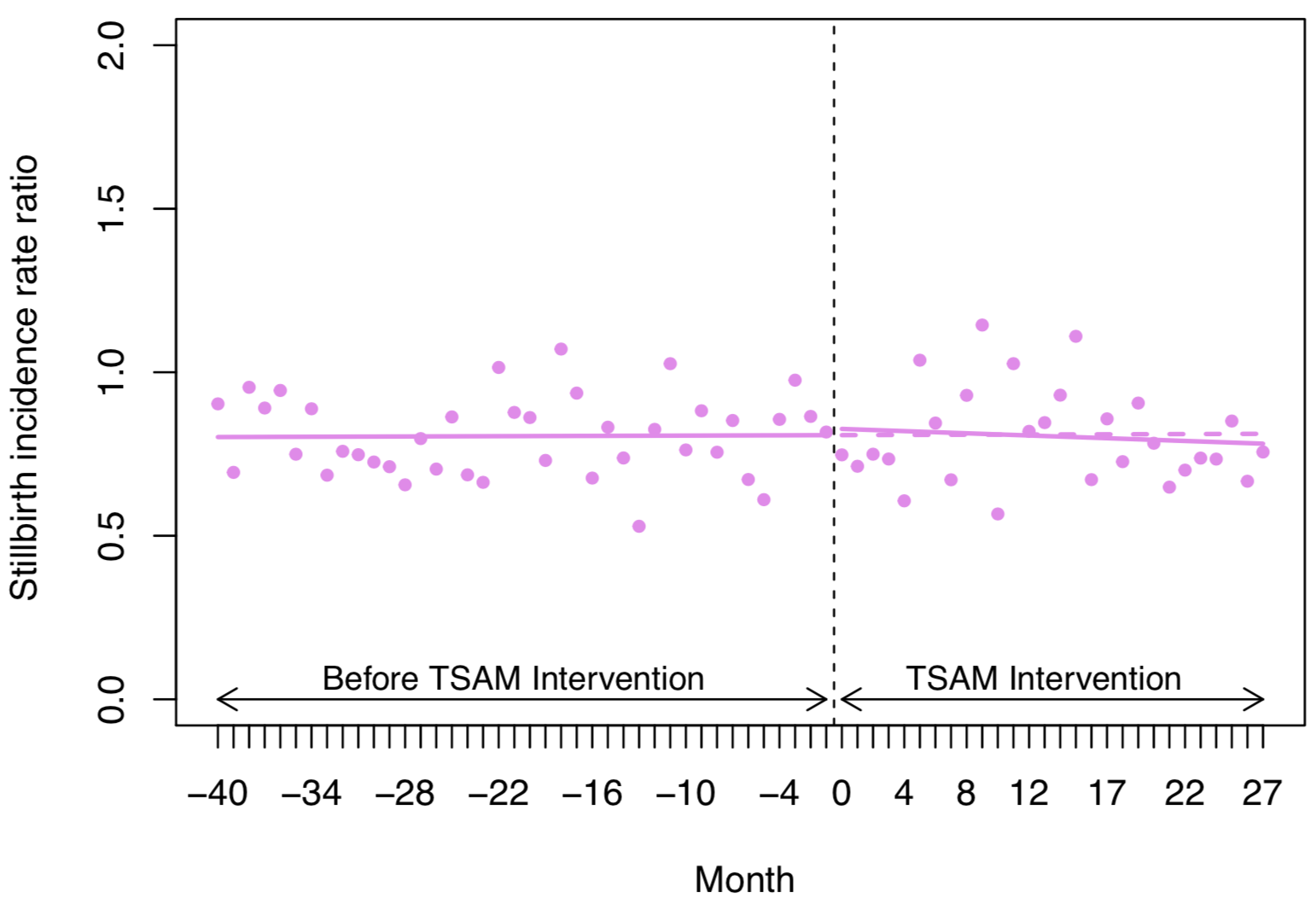

Appendix Figure 2. SITS analysis of impact of TSAM intervention on stillbirth rate ratio.

TSAM, training support and access model.

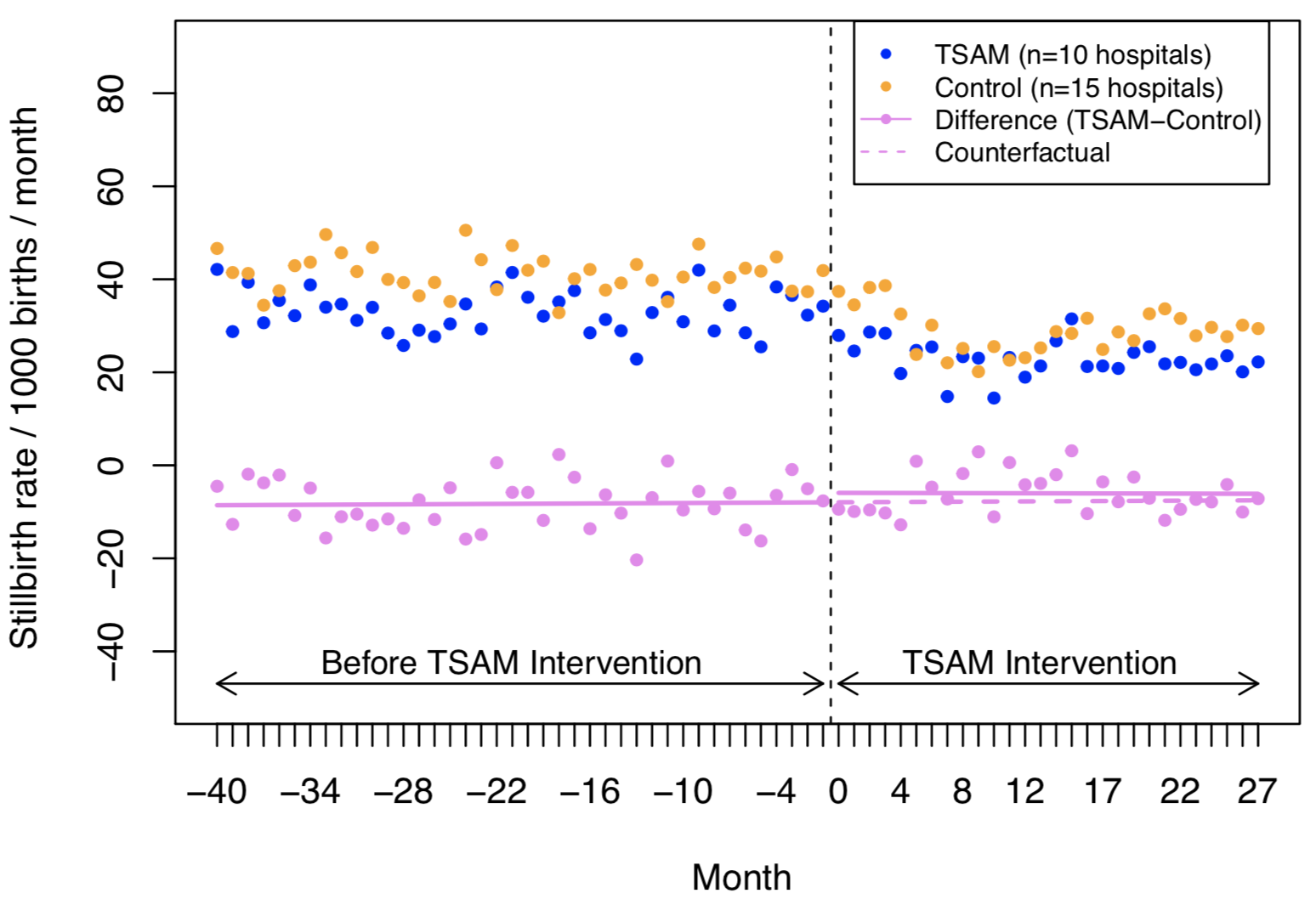

Appendix Figure 3. SITS analysis of impact of TSAM intervention on differences in stillbirth rate between intervention and control hospitals. TSAM, training support and access model.

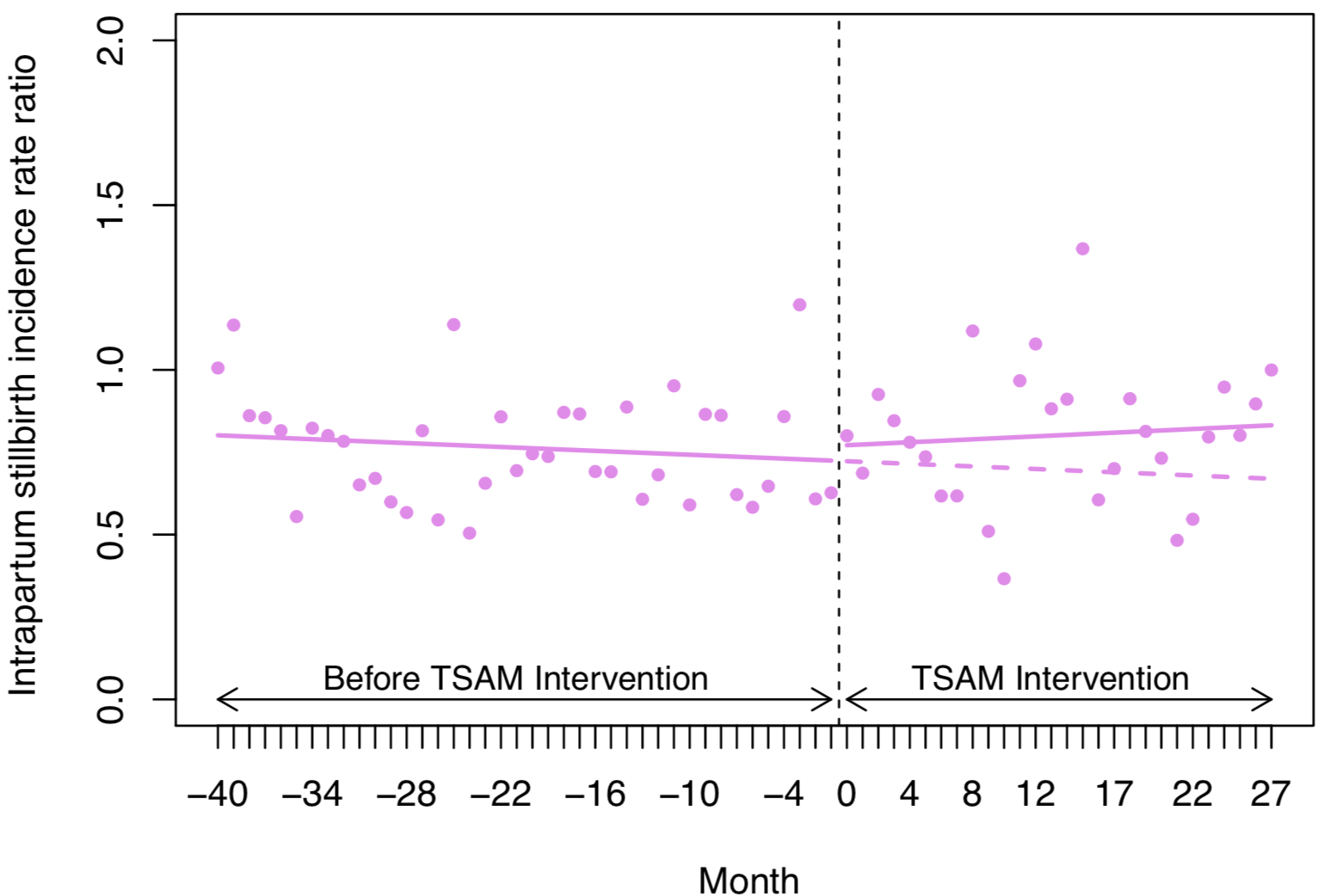

Appendix Figure 4. SITS analysis of impact of TSAM intervention on ratio of stillbirth rate between intervention and control hospitals.

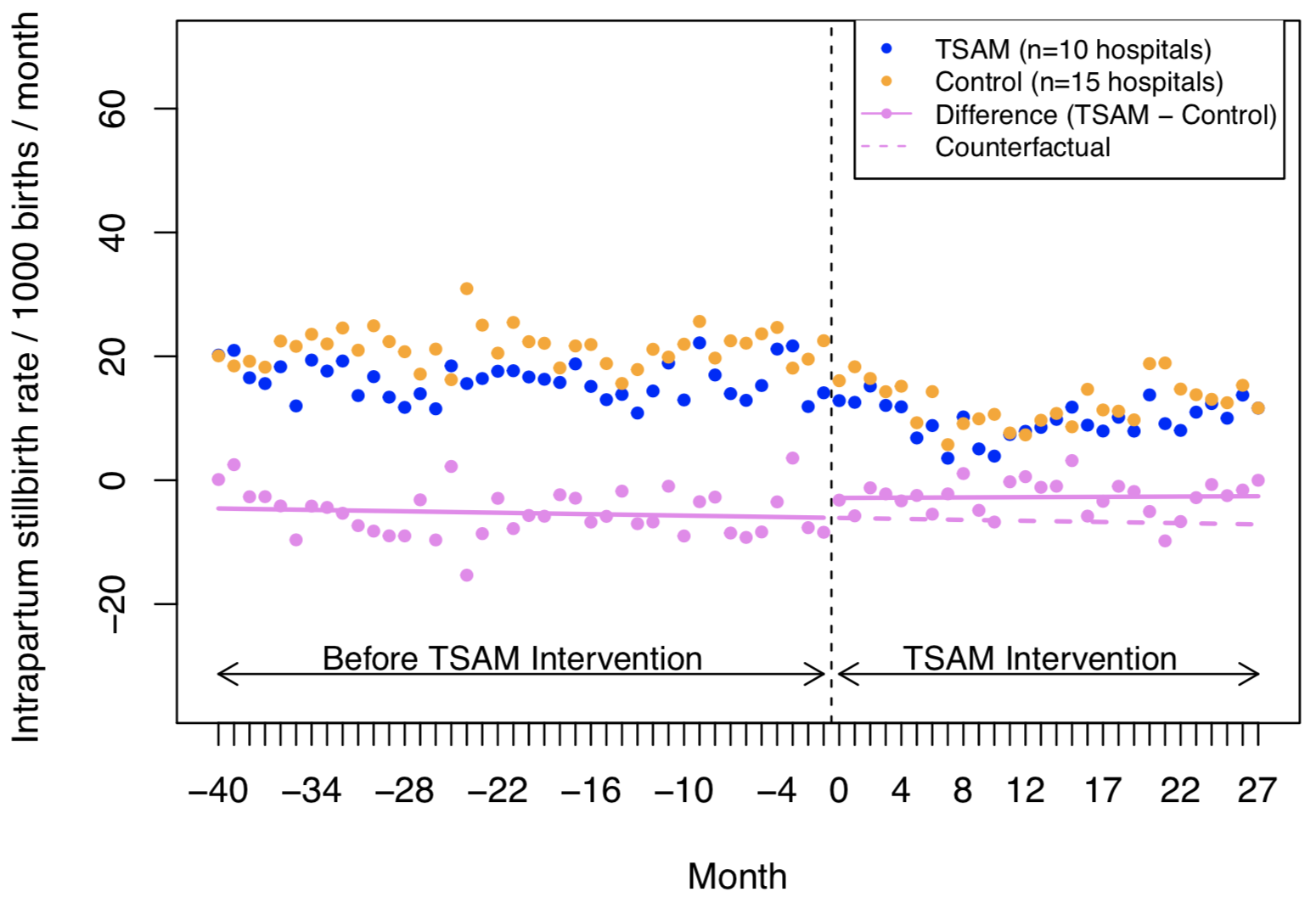

Appendix Figure 5. SITS analysis of impact of TSAM intervention on differences in stillbirth rate between intervention and control hospitals.

TSAM, training support and access model.

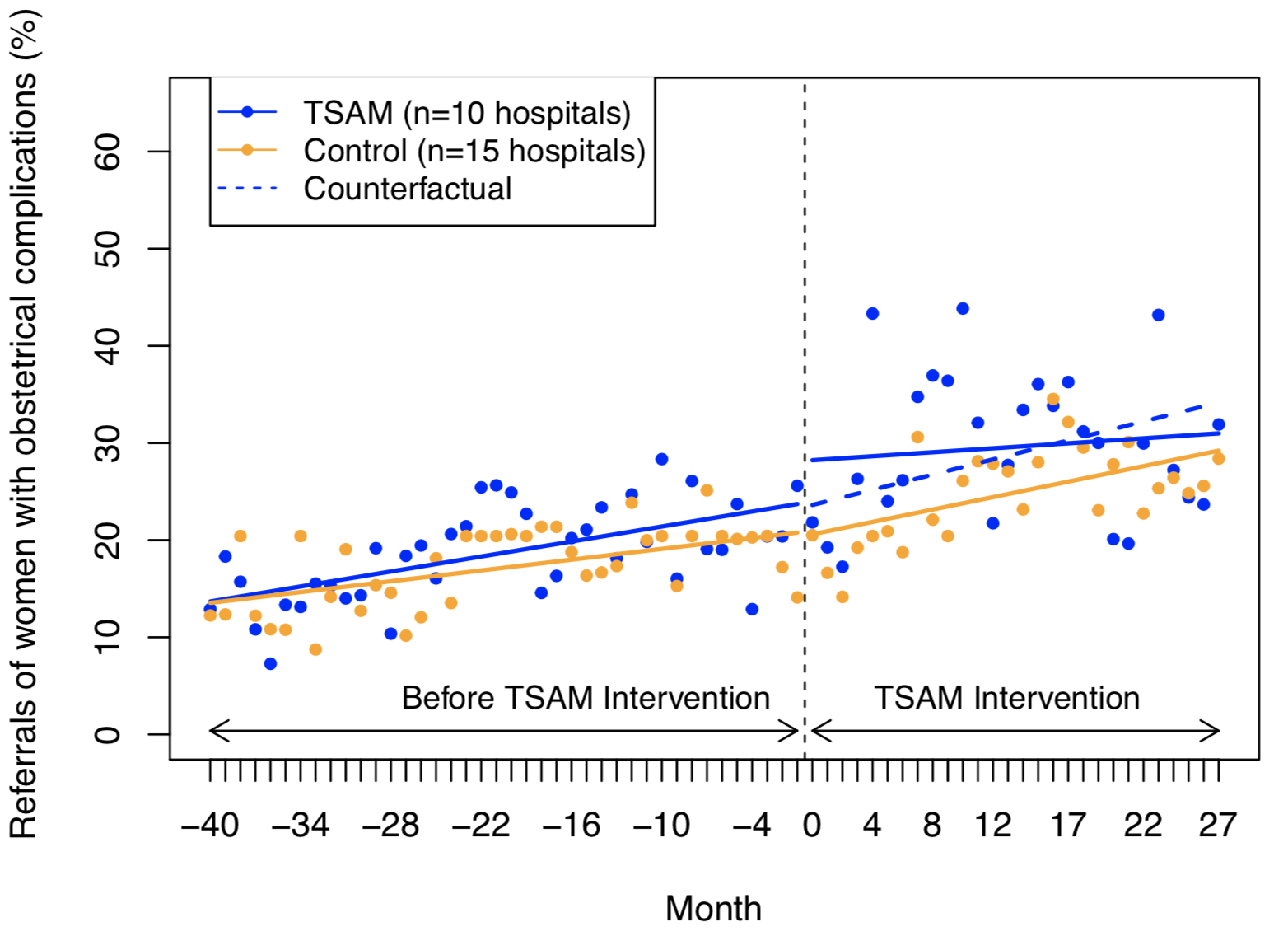

Appendix Figure 6. CITS analysis of impact of TSAM intervention on maternal referral rate to higher level of care.

TSAM, training support and access model.

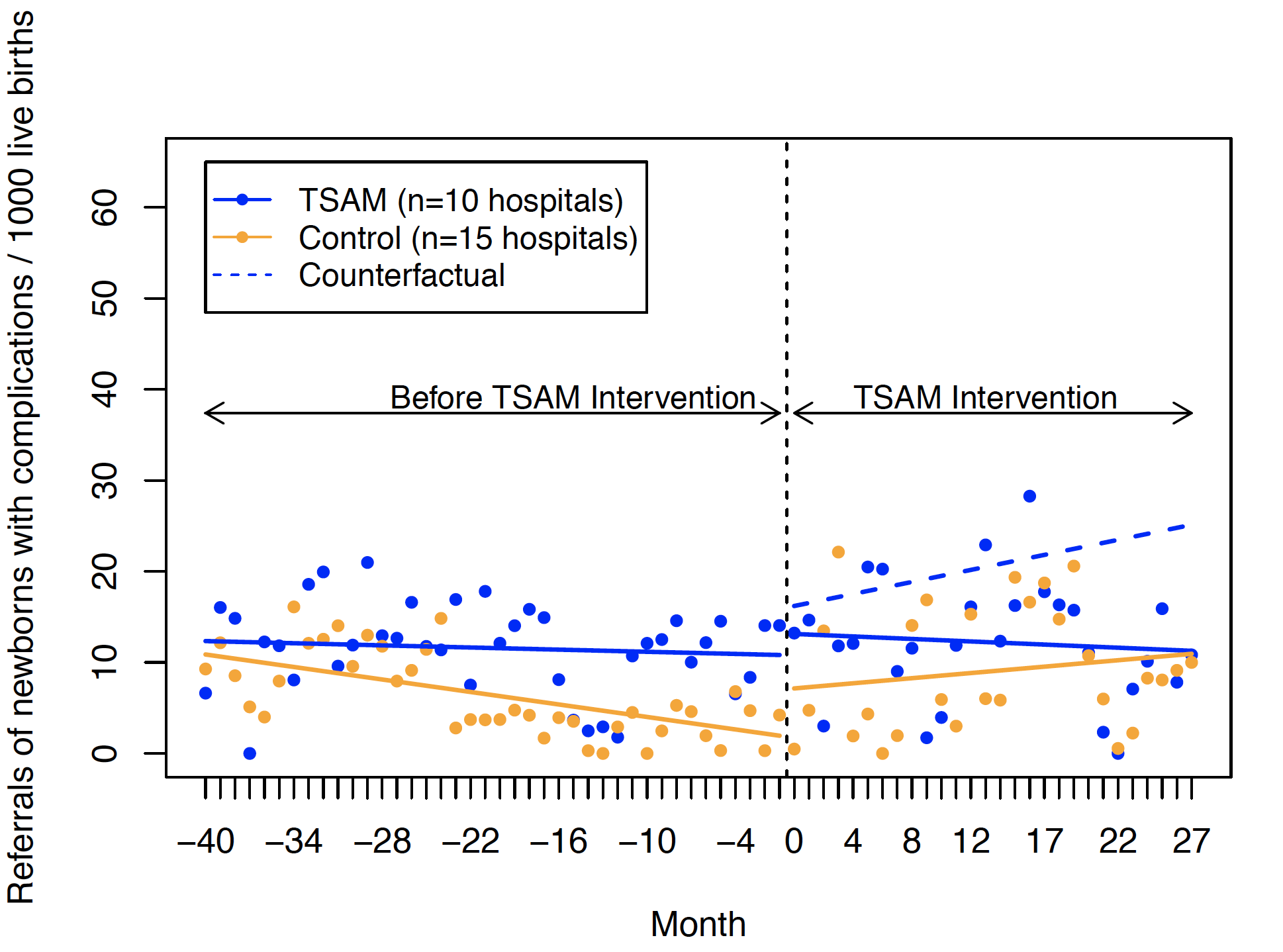

Appendix Figure 7. CITS analysis of impact of TSAM intervention on referral rate of newborns with complications at birth to higher level of care.

TSAM, training support and access model.

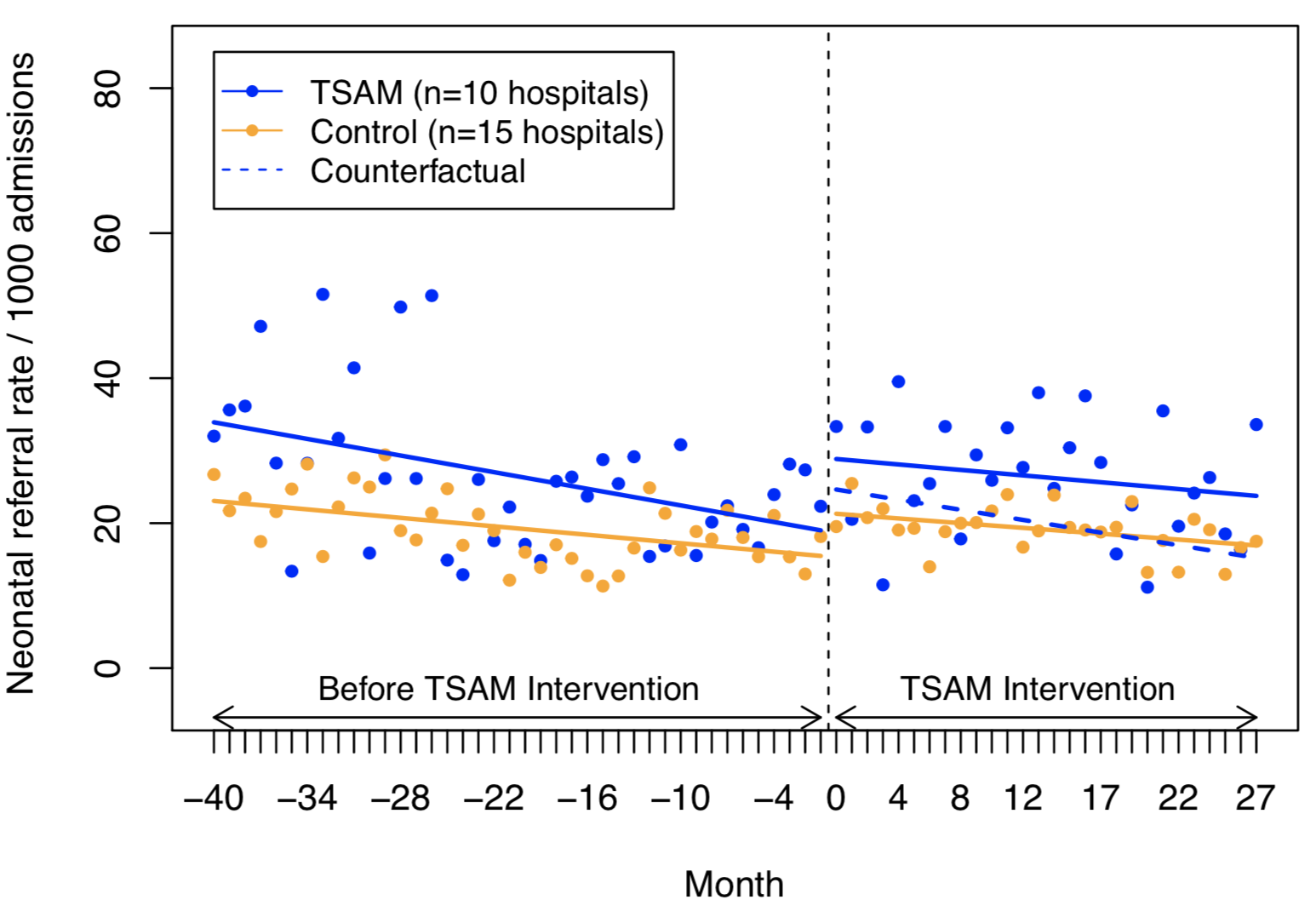

Appendix Figure 8. CITS analysis of impact of TSAM intervention on neonatal referral rate.

TSAM, training support and access model.

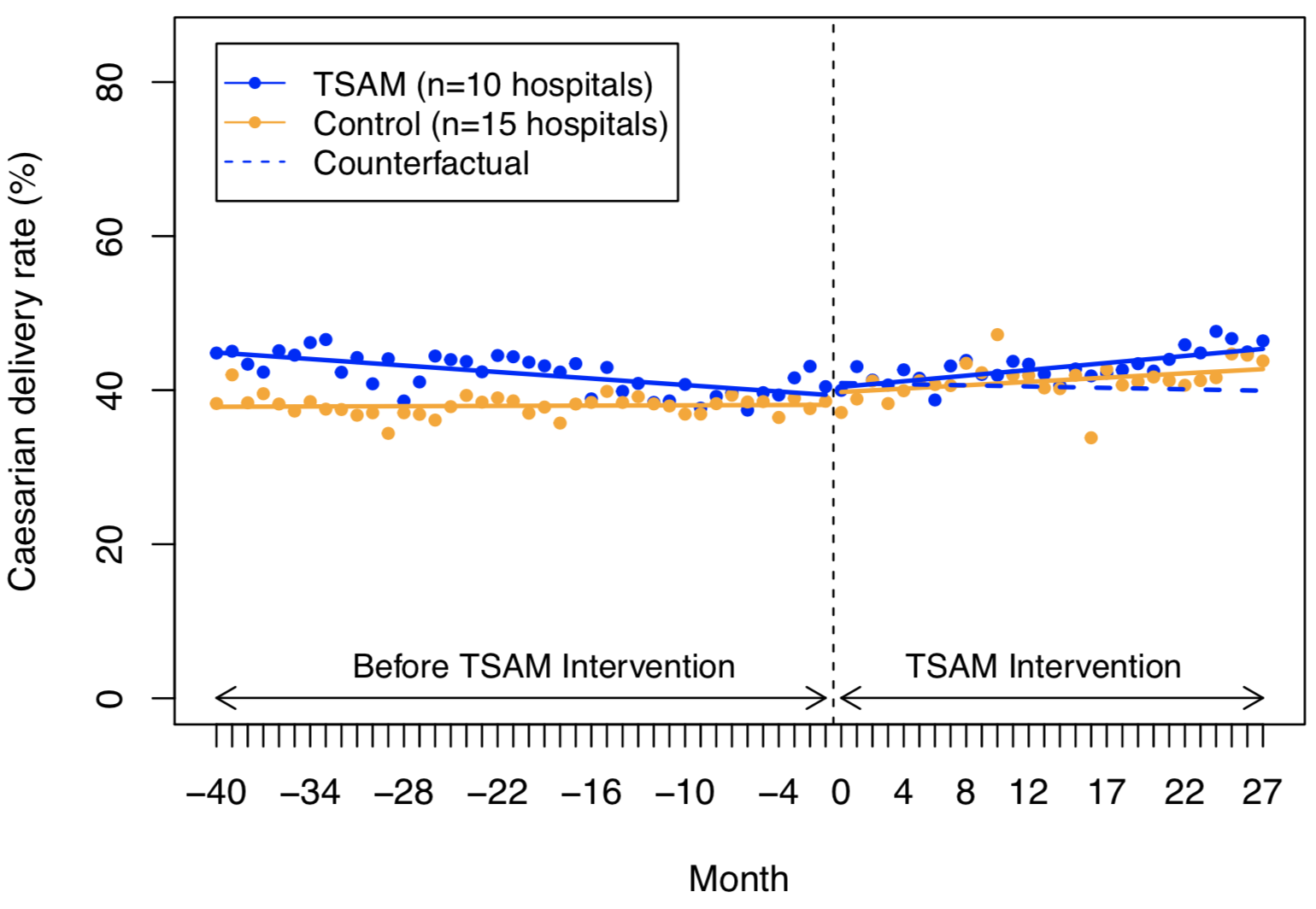

Appendix Figure 9. CITS analysis of impact of TSAM intervention on caesarian delivery rate.

TSAM, training support and access model.

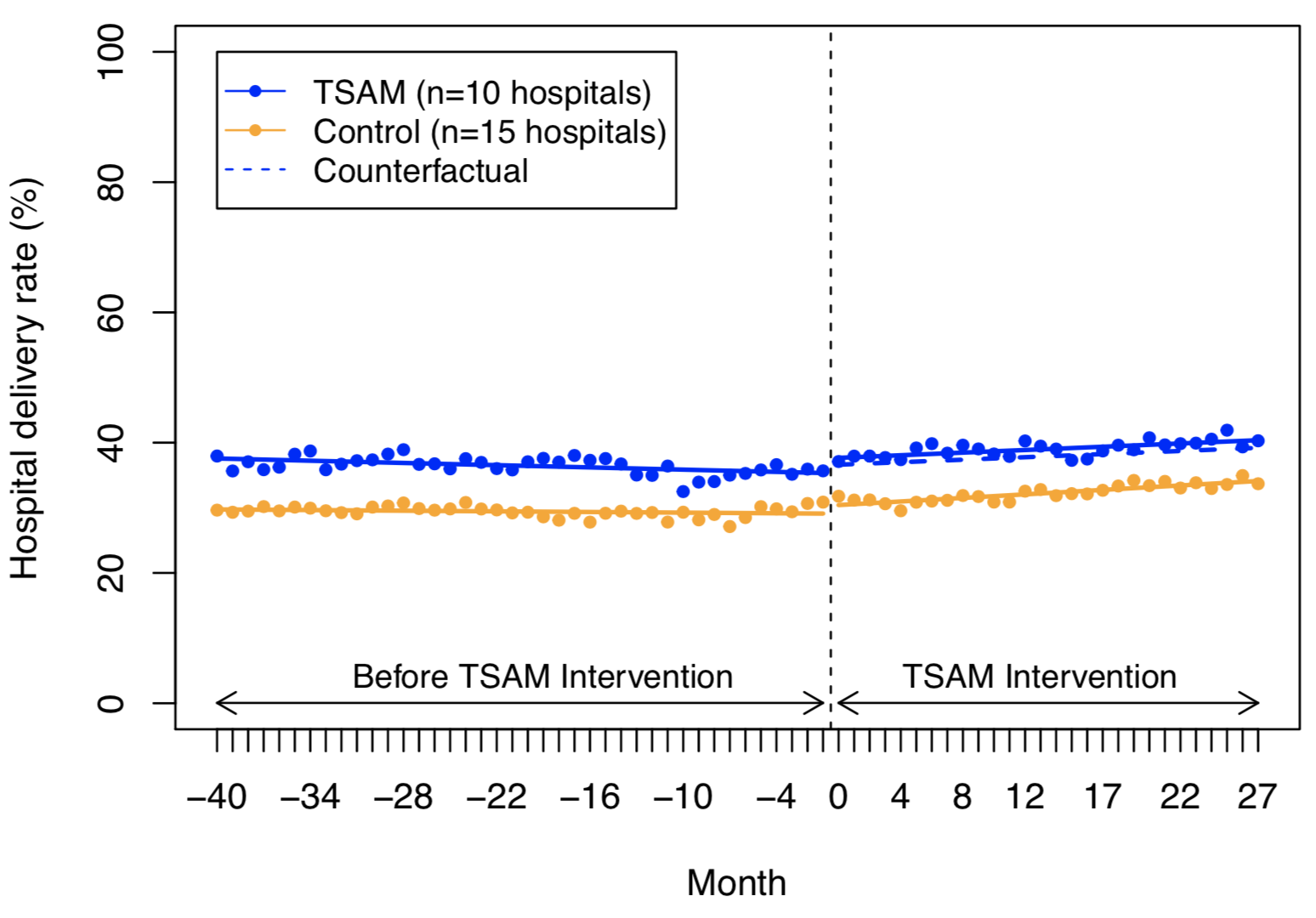

Appendix Figure 10. CITS analysis of impact of TSAM intervention on hospital delivery rate.

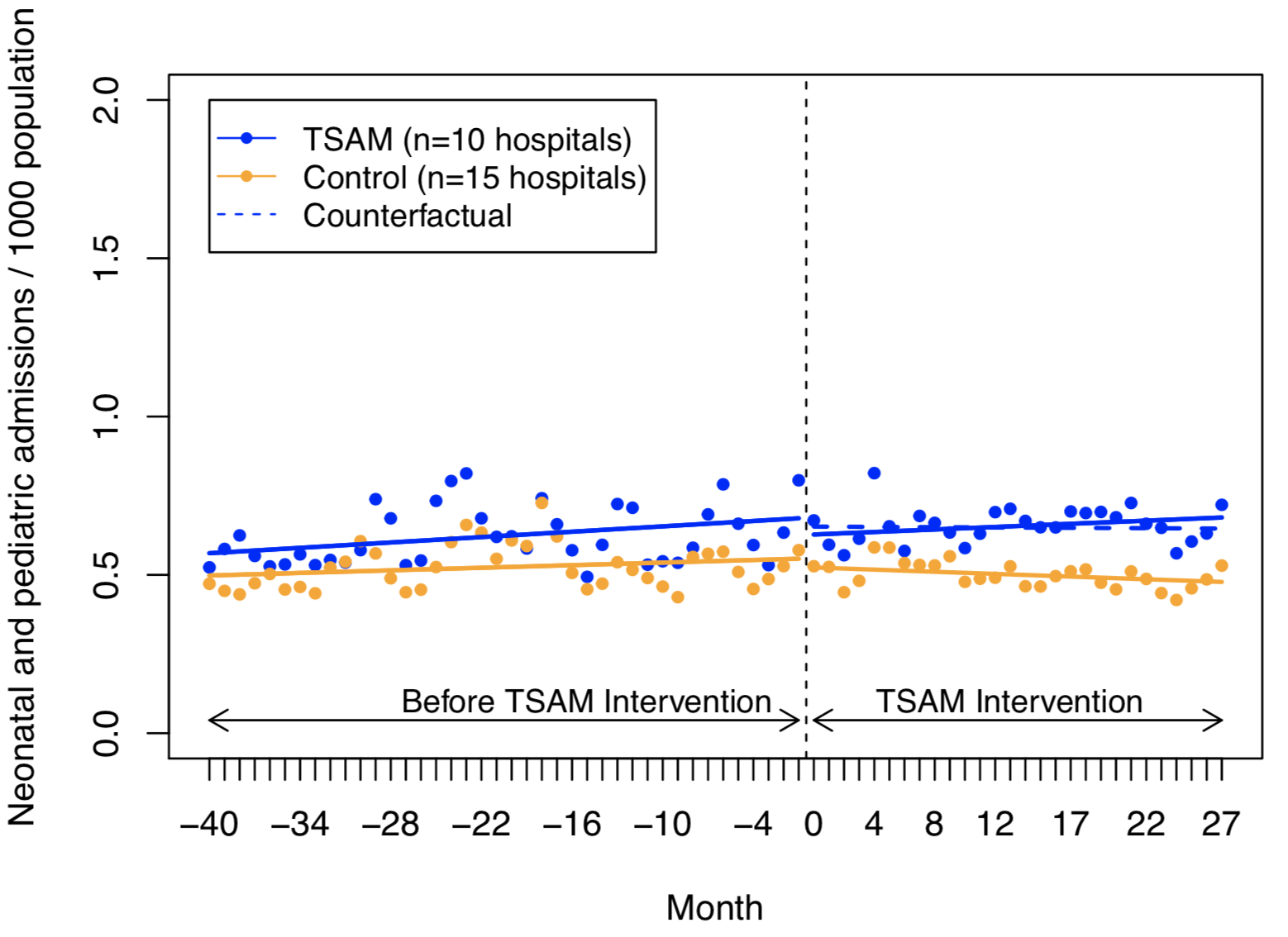

Appendix Figure 11. CITS analysis of impact of TSAM intervention on neonatal and pediatric admission rate.

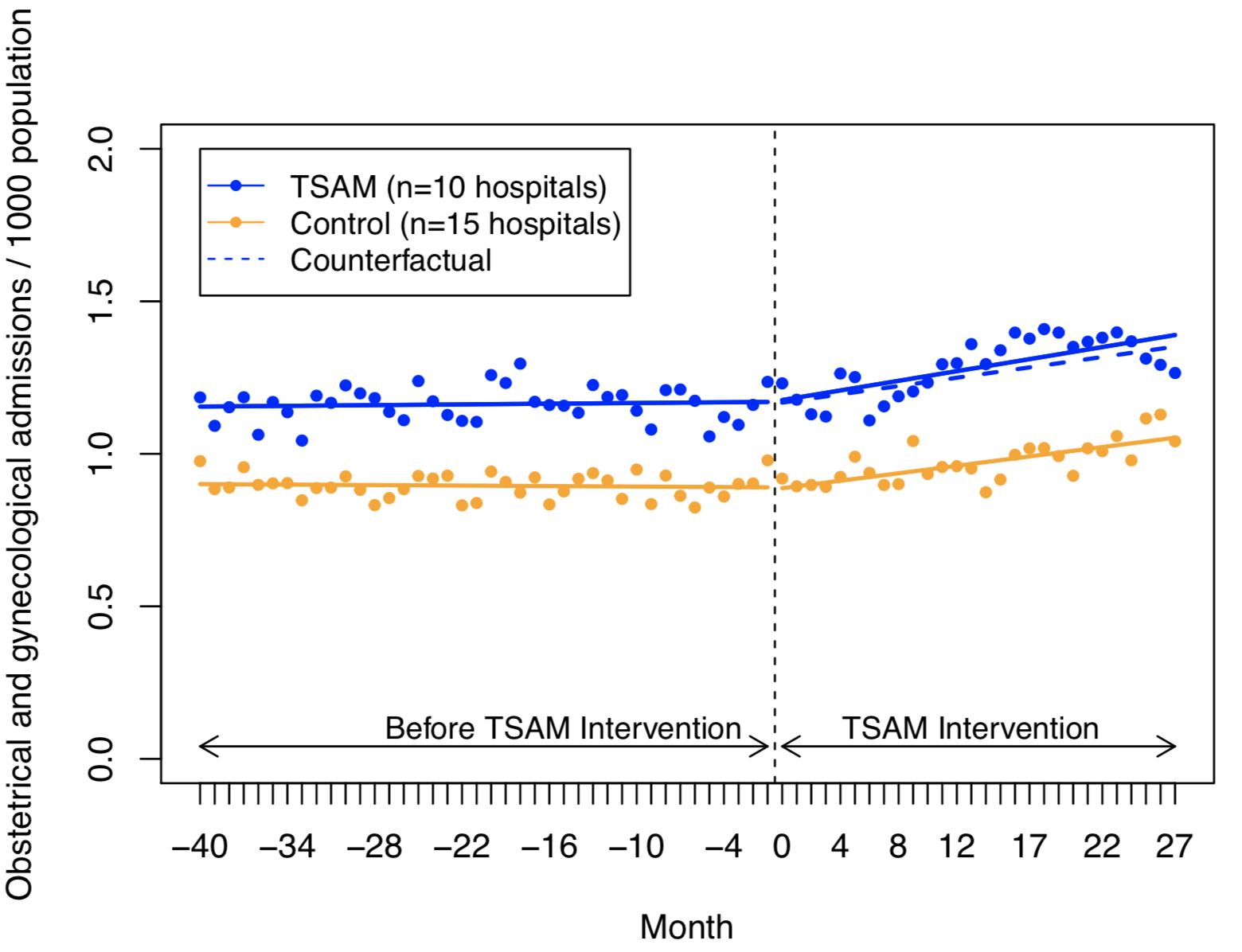

Appendix Figure 12. CITS analysis of impact of TSAM intervention on obstetrical and gynecological admission rate
